# Vaccines that prevent SARS-CoV-2 transmission may prevent or dampen a spring wave of COVID-19 cases and deaths in 2021

**DOI:** 10.1101/2020.12.13.20248120

**Authors:** David A. Swan, Ashish Goyal, Chloe Bracis, Mia Moore, Elizabeth Krantz, Elizabeth Brown, Fabian Cardozo-Ojeda, Daniel B Reeves, Fei Gao, Peter B. Gilbert, Lawrence Corey, Myron S. Cohen, Holly Janes, Dobromir Dimitrov, Joshua T. Schiffer

## Abstract

Ongoing SARS-CoV-2 vaccine trials assess vaccine efficacy against disease (VE_DIS_), the ability of a vaccine to block symptomatic COVID-19. They will only partially discriminate whether VE_DIS_ is mediated by preventing infection as defined by the detection of virus in the airways (vaccine efficacy against infection defined as VE_SUSC_), or by preventing symptoms despite breakthrough infection (vaccine efficacy against symptoms or VE_SYMP_). Vaccine efficacy against infectiousness (VE_INF_), defined as the decrease in secondary transmissions from infected vaccine recipients versus from infected placebo recipients, is also not being measured. Using mathematical modeling of data from King County Washington, we demonstrate that if the Moderna and Pfizer vaccines, which have observed VE_DIS_>90%, mediate VE_DIS_ predominately by complete protection against infection, then prevention of a fourth epidemic wave in the spring of 2021, and associated reduction of subsequent cases and deaths by 60%, is likely to occur assuming rapid enough vaccine roll out. If high VE_DIS_ is explained primarily by reduction in symptoms, then VE_INF_>50% will be necessary to prevent or limit the extent of this fourth epidemic wave. The potential added benefits of high VE_INF_ would be evident regardless of vaccine allocation strategy and would be enhanced if vaccine roll out rate is low or if available vaccines demonstrate waning immunity. Finally, we demonstrate that a 1.0 log vaccine-mediated reduction in average peak viral load might be sufficient to achieve VE_INF_=60% and that human challenge studies with 104 infected participants, or clinical trials in a university student population could estimate VE_SUSC_, VE_SYMP_ and VE_INF_ using viral load metrics.

## Introduction

The endpoint for SARS-CoV-2 vaccine efficacy trials targeting licensure is vaccine efficacy against disease (VE_DIS_) which is defined by a reduction in symptomatic disease, confirmed with polymerase chain reaction (PCR) testing for viral RNA, in vaccine recipients relative to placebo recipients (*1, 2*). The FDA benchmark for licensure is a point estimate of VE_DIS_>50% with lower alpha-adjusted 95% confidence limit exceeding 30% (*3*). Two mRNA vaccines have shown high levels of protection (>90%) at interim analyses (*4, 5*).

Once VE_DIS_ is established and a vaccine is licensed, mathematical modeling is useful for projecting a roll out strategy that affords maximal reductions in deaths and cases, and to prevent the need for future lockdowns (*6-8*). Yet VE_DIS_ does not provide sufficient information to fully inform these models. High VE_DIS_ is determined by a combination of two distinct phenomena which will only be partially captured in these trials: vaccine efficacy against susceptibility (VE_SUSC_) which is defined as the vaccine-induced reduction in the rate of infection – as evidenced by detection of virus by PCR - and vaccine efficacy against symptoms (VE_SYMP_) which is defined as the reduction in the presence of symptoms conditional on infection under vaccine versus placebo **(Table 1, Figure 1)** (*1, 2, 9*). If a vaccine mediates VE_DIS_ primarily through reduction in symptoms, the extent to which people, who convert from symptomatic to asymptomatic infection as a result of receiving the vaccine, can still transmit the virus, remains unknown. A vaccine that achieves high VE_DIS_ via VE_SYMP_ could theoretically contribute less to overall herd immunity than a vaccine that achieves high VE_DIS_ via VE_SUSC_, as the former may not block ongoing chains of transmission from vaccine recipients.

**Table 1.** Vaccine Efficacy definitions.

|  | **Definition** | **Formula** |
| --- | --- | --- |
| **Vaccine efficacy against symptomatic infection** | Reduction in virologically confirmed symptomatic COVID-19 in vaccine versus placebo recipients | $VE_{DIS} = 1 - (V_{DIS}/P_{DIS})$ |
| **Vaccine efficacy against infection** | Reduction in virologically confirmed asymptomatic or symptomatic SARS-CoV-2 infection in vaccine versus placebo recipients | $VE_{SUSC} = 1 - (V_{SUSC}/P_{SUSC})$ |
| **Vaccine efficacy against symptoms** | Reduction in development of symptoms conditional on infection in vaccine versus placebo recipients | $VE_{SYMP} = 1 - (V_{DIS}/V_{SUSC}) / (P_{DIS}/P_{SUSC})$ |
| **Vaccine efficacy against infection** | Reduction in number of secondary contacts infected by infected vaccine recipients versus number of secondary contacts infected by infected placebo recipients | $VE_{INF} = 1 - (V_{INF}/P_{INF})$ |

$V_{DIS}$ = % in the vaccine arm with virologically confirmed symptomatic COVID-19
$P_{DIS}$ = % in the placebo arm with virologically confirmed symptomatic COVID-19
$V_{SUSC}$ = % in the vaccine arm with virologically confirmed SARS-CoV-2 infection (symptomatic or asymptomatic)
$P_{SUSC}$ = % in the placebo arm with virologically confirmed SARS-CoV-2 infection (symptomatic or asymptomatic)
$V_{INF}$ = number of secondary contacts infected by vaccine recipients with virologically confirmed symptomatic COVID-19
$P_{INF}$ = number of secondary contacts infected by placebo recipients with virologically confirmed symptomatic COVID-19

**Figure 1.**
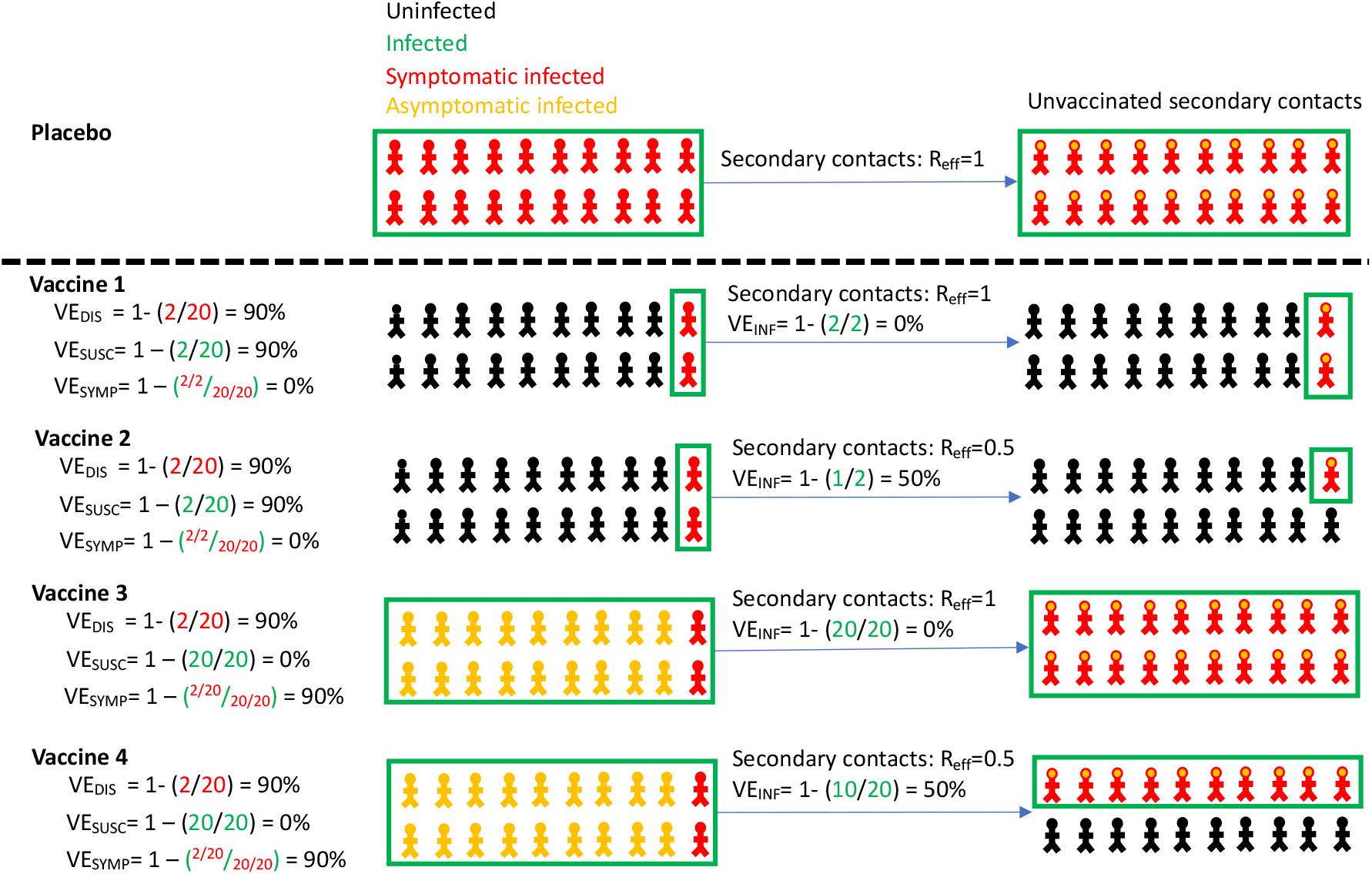
Vaccine efficacy definitions. Four vaccines with high efficacy against disease (VE_DIS_=90%) are demonstrated with different projected outcomes on vaccinated persons (left) and secondary contacts of infected person (right). Vaccines 1 and 2 mediate reduction of symptomatic infection by eliminating infection altogether, whereas vaccines 3 and 4 reduce symptoms among infected people. Vaccines 1 and 3 provide no reduction in secondary transmission risk. Vaccines 2 and 4 provide 50% reduction in secondary transmission risk. Definitions are in **Table 1**. All persons in the placebo arm are symptomatically infected for demonstration purposes only. Infected secondary contacts may be symptomatic or asymptomatic. R_eff_ is the effective reproductive number representing the number of secondary transmission per infected person.

A third vaccine effect, efficacy against infectiousness (VE_INF_) is defined as reduction in secondary transmissions from either symptomatic or asymptomatic infected vaccine versus placebo recipients and could also have significant effects on the trajectory of viral epidemics (*10*). Reduced VE_INF_ anticipates that symptomatic breakthrough infections in vaccine recipients may be associated with fewer secondary transmissions than in placebo recipients, and that people who develop asymptomatic rather than symptomatic infection due to vaccination (VE_SYMP_) may also be less likely to transmit. This latter observation would be expected if a vaccine mediates reduction in both symptoms and secondary transmission potential by lowering the quantity of viral shedding (*11*). While high VE_DIS_ guarantees a high likelihood of individual benefit, protection of unvaccinated members of the population will also depend on VE_SUSC_ and VE_INF_, as well as the velocity of a vaccination roll out program (*8, 12*).

SARS-CoV-2 serology is being used in the Pfizer and Moderna trials to capture asymptomatic infections and thereby estimate VE_SUSC_ and VE_SYMP_ (*1*). Yet, waning SARS-CoV-2 humoral responses could limit the sensitivity of this approach. Results from the ChAdOx1 vaccine trial show a trend towards lower protection against infection by viral nucleic acid detection than against symptomatic COVID-19, highlighting the potential importance of this approach, though low frequency of sampling could limit the accuracy and precision of these estimates (*13*). VE_INF_ is also not being directly assessed in ongoing vaccine trials. While all current studies will ultimately measure viral load at presentation among symptomatic infected persons only, this approach misses all asymptomatic people who may or may not continue to secondarily transmit SARS-CoV-2. It also does not capture the critical pre-symptomatic phase of symptomatic infection when viral load and transmissibility are highest (*14-16*).

The inability to fully discriminate VE_SUSC_ from VE_SYMP_, and to directly measure VE_INF_, in the current slate of promising vaccines limits our ability to forecast vaccine impacts in the population. Specifically, there is uncertainty regarding the threshold of vaccinated people required to achieve herd immunity, where the effective reproductive number (R_eff_) is maintained below 1 and new cases contract. It is similarly challenging to optimize vaccine allocation to different sectors of the population. For instance, it may be best to target a vaccine with high VE_SUSC_ or VE_INF_, which breaks secondary chains of transmission, towards essential workers and young people. Alternatively, a vaccine with high VE_SYMP_ but limited effects on secondary transmission may be best prioritized towards populations with highest risk of severe disease, such as the elderly (*8*).

Several possible methods exist to estimate VE_INF_. One is to measure secondary attack rate among household contacts of infected vaccine recipients versus infected placebo recipients (*17*). Alternatively, cluster randomized trials can assess for indirect protection of unvaccinated persons in vaccinated versus unvaccinated communities (*18*). While both of these trial designs are attractive, they have high operational complexity and will need to be implemented and completed rapidly to impact the course of the pandemic.

Another option is to use a viral load metric as a surrogate endpoint. VE_INF_ is likely to be mediated via a reduction in viral load among recipients of vaccine versus placebo, particularly early during pre-symptomatic or asymptomatic infection when nasal and saliva viral loads are highest (*16, 19-21*). It is possible that VE_SYMP_ is also driven by viral load reduction, though it has yet to be proven beyond association whether any specific viral load metric predicts development of symptoms or severe COVID-19 (*22*). Moreover, only a few studies captured critical early peak viral load kinetics, and in too few people to perform correlate analyses (*20*). Viral load in infected vaccine versus placebo recipients could be measured in large clinical trials in which enrolled participants undergo frequent self-sampling after enrollment, or in smaller highly controlled human challenge studies (*23*).

Here we use a mathematical modeling approach using data from King County Washington to demonstrate the potential effects of VE_INF_ at the population level given multiple vaccine profiles. In contrast to existing models of vaccine prioritization (*7, 8*), including our own (*24*), the model accounts for the likely need for recurrent lockdowns once cases and hospitalizations exceed a certain threshold. We next estimate reduction in peak viral load required to achieve various VE_INF_ and outline animal viral graded challenge experiments required to verify the relationship between viral load and likelihood of transmission. Finally, we propose human challenge study design, and describe a potential clinical trial in university students, to rapidly estimate VE_SUSC_, VE_SYMP_ and VE_INF_ for a vaccine.

## Results

### Overview

We use a mathematical model of COVID-19 in King County Washington to project the impact of different vaccine profiles on incident cases, hospitalizations and deaths throughout 2021. Our goal is to identify vaccine efficacy profiles under which high VE_INF_ would or would not reduce a large number of infections and deaths, as well as the need for prolonged lockdowns. Because we identify that VE_INF_ could determine whether or not a large fourth wave of infections occurs in the region during the spring of 2021, and because VE_INF_ has not yet been measured for vaccines under study, we next focus on approaches to rapidly estimate its value. Using an intra-host transmission model (*14, 25, 26*), we consider reduction in peak viral load as a potential surrogate endpoint for VE_INF_, discuss graded challenge studies in animal models to validate these predictions, and provide human viral challenge study designs which might provide actionable VE_INF_ estimates within relevant timeframes for the pandemic.

### Vaccine efficacy definitions

We include vaccines defined by four different types of efficacy **(Table 1)**. VE_DIS_ is the primary endpoint in all ongoing phase 3 vaccine trials and is defined as the proportion of vaccine versus placebo recipients who do not develop symptomatic COVID-19 (*1*). VE_DIS_ represents a composite of efficacy against infection (VE_SUSC_) and disease (VE_SYMP_). VE_SUSC_ is the proportion of people in a vaccine arm relative to the placebo arm of a trial who are fully protected against both asymptomatic and symptomatic virologically confirmed SARS-CoV-2 infection. The effect of high VE_SUSC_ (90%) is shown in **Vaccines 1 & 2** in **Fig 1**. Complete protection against infection also implies no possibility of secondary transmission. VE_SYMP_ is conditional on infection and is defined as the proportion of people in the vaccine arm relative to the placebo arm of a trial who remain asymptomatic despite being infected. A vaccine in which VE_DIS_ is mediated entirely by VE_SYMP_ will not lower the absolute number of people infected **(Vaccines 3 & 4** in **Fig 1**).

The proportion of secondary contacts who are symptomatically or asymptomatically infected (red and orange figures respectively in **Fig 1**) by infected vaccine recipients relative to secondary contacts of placebo recipients determines the vaccine efficacy against infectiousness (VE_INF_, **Table 1**). Unlike VE_SUSC_ and VE_SYMP_, VE_INF_ is not a determinant of VE_DIS_. Vaccines with no VE_INF_ **(Vaccines 1 & 3**) and moderate VE_INF_ **(Vaccines 2 & 4**) are shown in **Fig 1**. Notably, the addition of VE_INF_=50% prevents a much larger number of secondary infections in a scenario where high VE_DIS_ is mediated entirely by VE_SYMP_ **(Vaccine 4)** versus a scenario where high VE_DIS_ is mediated entirely by VE_SUSC_ **(Vaccine 2)**.

The possible relevance of this concept for the Moderna and Pfizer vaccines is evident by comparing extreme scenarios in which VE_DIS_=90%. A vaccine with VE_DIS_=90% due to VE_SUSC_=90% / VE_SYMP_=0% / VE_INF_=0% would have very similar population level effects as a vaccine with VE_SUSC_=0% / VE_SYMP_=90% / VE_INF_=100%. In the latter scenario, all vaccinated and subsequently infected people would be asymptomatic and could also not secondarily transmit the virus. On the other hand, a vaccine with VE_SUSC_=0% / VE_SYMP_=90% / VE_INF_=0% would protect vaccinated people against symptoms but have no impact on downstream transmissions.

### Projection of incident SARS-CoV-2 infections and deaths in King County Washington in 2021 assuming no vaccine

We modified a previously developed compartmental model to reproduce the ongoing COVID-19 epidemic in King County Washington (*24*). The model includes uninfected, exposed, asymptomatic infected, pre-symptomatic infected, symptomatic infected, diagnosed asymptomatic, diagnosed symptomatic, hospitalized, dead and recovered compartments, all stratified into four age cohorts **(Sup fig 1)**. We calibrated the model to daily cases **(Sup fig 2a)**, daily hospitalizations **(Sup fig 2b)**, daily deaths **(Sup fig 2c**), age-stratified cases **(Sup fig2d)**, age-stratified hospitalizations **(Sup fig2e)**, age-stratified deaths **(Sup fig2f)**, cumulative cases **(Sup fig 2g)**, and cumulative deaths **(Sup fig 2h)** through November, 2020. Model equations are in the **Supplement** with parameters and their values in **Supplementary tables 1-4**.

Extending beyond the calibration period, we attempt to capture realistic approximations of local pandemic management to date. First, based on experience in other U.S. states and current Washington state policies, we assume that in the absence of a vaccine, numbers of cases and hospitalizations are likely to fluctuate due to the community response to the epidemic (*27, 28*). When number of new infections remains below a certain threshold, physical distancing measures are assumed to relax allowing greater contact between susceptible and infected people. The effective reproductive number (R_e_) eventually exceeds one and cases start growing in number. Eventually a threshold is surpassed that necessitates reinforcement of physical distancing restrictions: R_e_ drops below one and cases contract. Based on timing of Washington state reinforcement of social distancing (*29*), we set this threshold at a 2-week daily case average of 300 new cases per 100,000. Physical distancing is set on a scale of 0 to 1 where 0 represents the level of pre-pandemic interactivity in the population and 1 implies no physical interactions.

Without a vaccine (black line, **Fig 2**), we projected the ongoing recurrent third wave (numbered in top row) of infections **(Fig 2a)**, diagnosed cases **(Fig 2b)**, hospitalizations **(Fig 2c)** and deaths **(Fig 2d)** between 11/2020 and 3/2021 **(Fig 2, top row)**. We also anticipated the need to re-enforce physical distancing to achieve 40% interactivity relative to pre-pandemic levels **(Fig 2e)**, in order to lower R_e_ below 1 **(Fig 2f)**. The model projects that this third wave would have a significantly higher number of infections, diagnosed cases and hospitalizations then the first wave in the spring of 2020 and the second wave in the summer of 2020. Daily deaths were projected to peak at similar levels to the first wave due to a higher proportion of younger individuals becoming infected with lower death rates. By January 2021, despite physical distancing, ∼15% of the population was projected to have been infected with ∼7000 hospitalizations and ∼1500 deaths **(Fig 2, middle row**).

**Figure 2.**
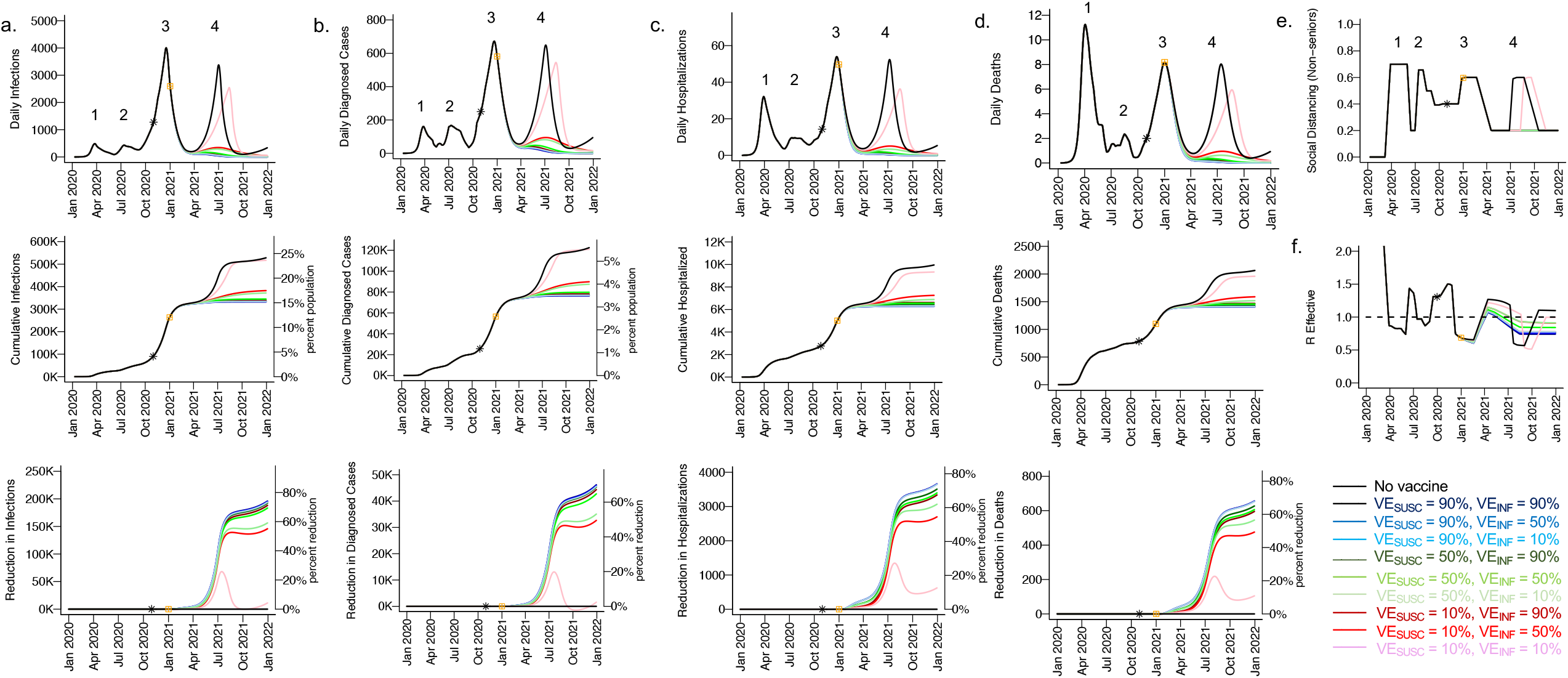
High VE_SUSC_ or high VE_INF_ alone can effectively limit cases and deaths with initial vaccine prioritization to the elderly. For unvaccinated (black lines) and each vaccine cohort red lines, legend), we project **a**. infections, **b**. diagnosed cases, **c**. hospitalizations and **d**. deaths, as well as **e**. social distancing relative to pre-pandemic levels and **f**. the effective reproductive number. The first four columns **(a-d)** are organized by row: top = daily incidence, middle = cumulative, bottom = reduction since day of vaccination. Waves of infection are numbered 1-4. Nine combinations of VE_SUSC_ and VE_INF_ are considered while VE_SYMP_ is fixed at 10%. High VE_SUSC_ (90%) simulations are blue and have similar outcomes to one another. Moderate VE_SUSC_ (50%) simulations are green. Low VE_SUSC_ (10%) simulations are red / pink. Dark lines are high VE_INF_ (90%) and have similar outcomes to one another. Moderate darkness are medium VE_INF_ (50%). Light lines are low VE_INF_ (10%). The largest reduction in cases is associated with either high VE_SUSC_ or VE_INF_. 5000 vaccines are given per day starting January 21 (yellow square) until 50% are vaccinated. Case threshold for reinstituting physical distancing to 0.6 is 300 per 100,000 and for relaxation is 100 per 100,000. 80% of vaccines are initially allocated to the elderly with the remaining 20% to middle-aged cohorts.

In the absence of a vaccine, our model projected a substantial fourth wave of infections **(Fig 2a)**, diagnosed cases **(Fig 2b)**, hospitalizations **(Fig 2c)** and deaths **(Fig 2d)** between April and October 2021, necessitating a fourth cycle of increased physical distancing **(Fig 2e)**. At the end of this fourth wave, we forecast that ∼25% of the population will have been infected and ∼5% diagnosed with ∼10,000 hospitalizations and ∼2000 deaths **(Fig 2, middle row**).

### Vaccine model scenarios

We consider scenarios in which VE_SUSC_, VE_SYMP_, and VE_INF_ each have either low (10%), medium (50%) or high (90%) efficacy. Each possible parameter combination allows for 3^3^ (27) vaccine scenarios. Five VE_SUSC_ and VE_SYMP_ combinations (15 scenarios when considering the 3 values of VE_INF_): VE_SUSC_=90% / VE_SYMP_=10%; VE_SUSC_=10% / VE_SYMP_=90%; VE_SUSC_=90% / VE_SYMP_=50%; VE_SUSC_=50% / VE_SYMP_=90%; VE_SUSC_=90% / VE_SYMP_=90%) would be roughly compatible with current projections for the Moderna and Pfizer mRNA vaccines which had estimated VE_DIS_ = 95% and 90% respectively at interim analyses (*4, 5*). Three combinations or 9 scenarios (VE_SUSC_=50% / VE_SYMP_=50%; VE_SUSC_=50% / VE_SYMP_=10%; VE_SUSC_=10% / VE_SYMP_=50%) would be realistic if there is a slight decrease in VE_DIS_ in the final analysis of these studies and would still meet criteria for licensure. These lower vaccine estimates may be relevant for other vaccines in development, or after a single dose of the Moderna or Pfizer product. One combination or 3 scenarios (VE_SUSC_=10% / VE_SYMP_=10%) which would not meet licensure requirements are included as controls to independently assess the effect of increasing VE_INF_.

We initially assumed 5000 vaccinations per day with the goal of covering 50% of the population of 2.2 million people with a start day for vaccination on January 1, 2021 allowing 220 days until completion in mid-August. In our simulations, both susceptible and recovered persons were vaccine eligible. We assumed that the vaccine start date represented timing of the second shot for the mRNA vaccines such that efficacy accrues at the defined time of vaccination. Initially, we imputed no loss of vaccine efficacy over time.

In one scenario **(Fig 2)**, we assumed disproportionate initial targeting of the cohorts aged > 70 initially (80% of vaccines with 20% to those older than 20 years old). In a second scenario **(Sup fig 3)**, we first vaccinated the age cohorts with greatest inter-connectivity (20-45 and 45-69 years old received 80% of vaccines). We also imputed slow relaxation of social distancing during the vaccination program when cases remained below a certain threshold.

### High VE_SUSC_ without VE_INF_ is sufficient for prevention of a fourth wave of SARS-CoV-2 cases, deaths, and lockdown in spring 2021

We first considered scenarios in which elderly cohorts were vaccinated first and VE_DIS_ was mediated mostly by VE_SUSC_ rather than VE_SYMP_ (VE_SYMP_=10%). Vaccines with high VE_SUSC_ (90%, blue lines) or VE_INF_ (90% darkest blue, green and red lines) resulted in the greatest reduction in peak **(Fig 2 top row)** and cumulative **(Fig 2 middle row)** infections **(Fig 2a)**, diagnosed cases **(Fig 2b)**, hospitalizations **(Fig 2c)** and deaths **(Fig 2d)**. All vaccines with VE_SUSC_ = 50% (green lines) or 90% (blue lines), or VE_INF_ = 50% (medium blue, green and red) or 90% (dark blue, green and red), prevented a large fourth wave of infections and deaths and allowed 20% physical distancing starting in April 2021 **(Fig 2e)** while maintaining R_e_ less than 1 **(Fig 2f)**. Notably, only VE_SUSC_=90% vaccines (blue lines) are compatible with Moderna and Pfizer results and VE_INF_ had little impact on these projections.

All vaccines with at least 90% VE_SUSC_ or VE_INF_, or both 50% VE_SUSC_ and 50% VE_INF_, lead to a reduction in approximately 200,000 infections, 45,000 diagnosed infections, 3500 hospitalizations and 600 deaths since the start of the vaccination period **(Fig 2 bottom row)**, though fewer deaths were prevented than would have already occurred by April 2021 (∼1500). A vaccine with VE_SUSC_ = 10%, VE_INF_ = 10% and VE_SYMP_ = 10% (pink line) was predicted to only delay the peak of infections, hospitalizations and deaths **(Fig 2 top row)** with a small percentage reduction in these outcomes over time **(Fig 2 bottom row)** and a requirement for a fourth phase of increased physical distancing **(Fig 2e)**.

Nearly equivalent results were noted when younger and middle-aged cohorts with higher inter-connectivity were vaccinated with higher priority **(Sup fig 3)**.

**Figure 3.**
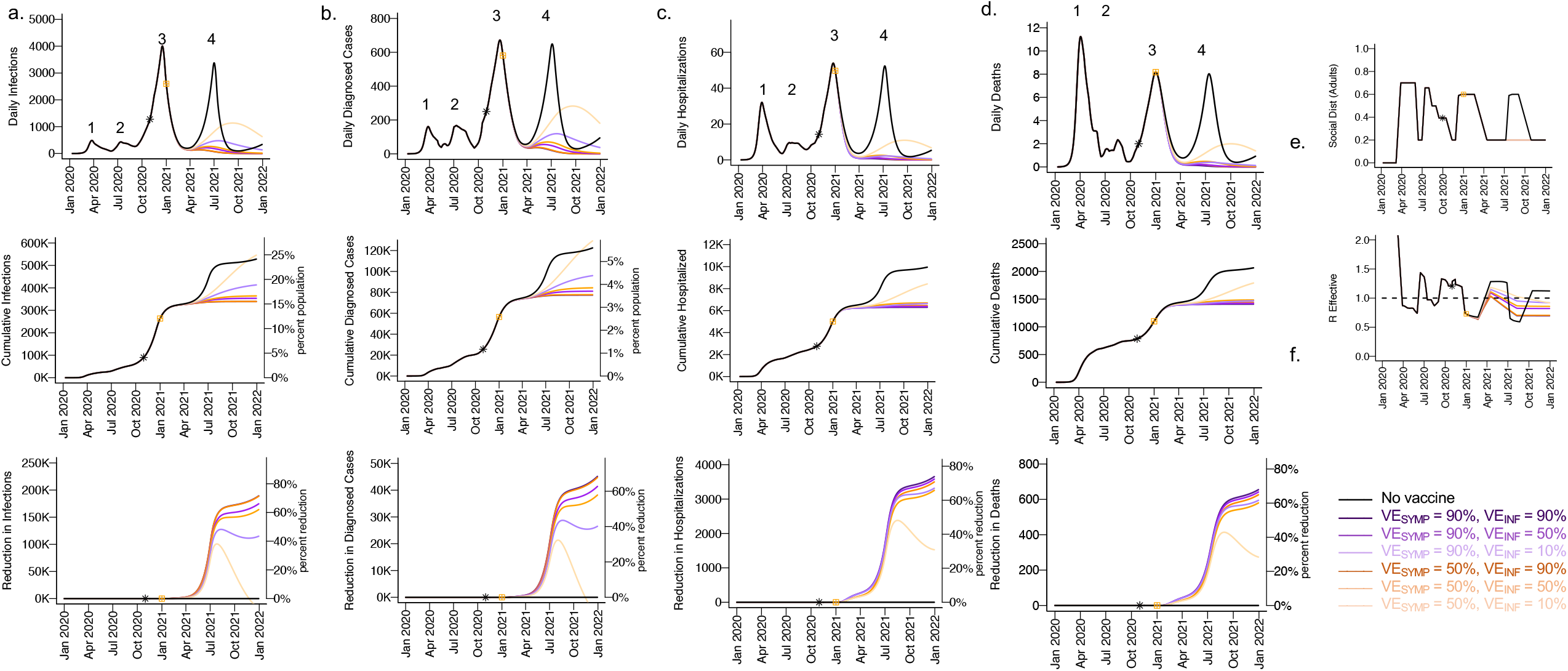
High VE_SYMP_ alone results in only partial reduction in infections given initial vaccine prioritization to the elderly population. For unvaccinated (black lines) and each vaccine cohort (colored lines), we project **a**. infections, **b**. diagnosed cases, **c**. hospitalizations and **d**. deaths, as well as **e**. physical distancing relative to pre-pandemic levels and **f**. the effective reproductive number. The first four columns (**a-d**) are organized by row: top = daily incidence, middle = cumulative, bottom = reduction since day of vaccination. Waves of infection are numbered 1-4. Six combinations of VE_SYMP_ and VE_INF_ are considered while VE_SUSC_ is fixed at low 10%. High VE_SYMP_ (90%) simulations are purple. Moderate VE_SYMP_ (50%) simulations are e. Dark lines are high VE_INF_ (90%). Moderate darkness lines are medium VE_INF_ (50%). Light lines are low VE_INF_ (10%). The largest reduction in cases is associated with high VE_INF_. vaccines are given per day starting January 1, 2021 (yellow square) until 50% are vaccinated. Case threshold for reinstituting physical distancing to 0.6 is 200 per 100,000 and for relaxation is 100 per 100,000.

### High VE_INF_ is a requirement for prevention of a fourth wave of SARS-CoV-2 cases, deaths, and lockdown in spring 2021 for low VE_SUSC_, high VE_SYMP_ vaccines

We next considered a scenario in which VE_DIS_ was mediated mostly by VE_SYMP_ rather than VE_SUSC_ (10%) with vaccine prioritization to the elderly. Once again, for all conditions with VE_INF_ = 90% (darkest purple, darkest orange lines), we observed a substantial decrease in infections **(Fig 3a)**, diagnosed cases **(Fig 3b)**, hospitalizations **(Fig 3c)** and deaths **(Fig 3d)** with no large fourth wave observed **(Fig 3, top row)**, a levelling off in cumulative incidence of all outcomes **(Fig 3, middle row)**, and a 60-70% reduction in all outcomes starting at the time of vaccination **(Fig 3, bottom row)**. There were no visible effects of increasing VE_SYMP_ when VE_INF_ = 90%.

Under the high VE_SYMP_, low VE_INF_ scenario compatible with the Moderna and Pfizer vaccine trial results (light purple line), a fourth protracted wave of ∼60,000 infections and ∼200 deaths lasting from April 2021 through January 2022 occurred which did not meet a threshold that necessitated further resumption of lockdown measures **(Fig 3e**,**f)**. At moderate VE_INF_ (50%, medium purple and medium orange) and in particular at lower VE_INF_ (10%, light purple and light orange), we observed a beneficial effect of increased VE_SYMP_ **(Fig 3)** with further reduction in all outcomes at high (90%, light purple) versus moderate (50%, light orange) VE_SYMP_. This result likely relates to the fact that VE_SYMP_ converts symptomatic to asymptomatic infection which in our model is predicted to be associated with a 44% decrease in overall infectivity.

### Lowering of cases and deaths under vaccine scenarios with low or moderate VE_SUSC_ and high VE_INF_

We explored the impact of varying VE_INF_ under several plausible vaccine scenarios with VE_DIS_ = 90%. We generated heat maps for total post-vaccine diagnosed cases **(Fig 4a)** and deaths **(Fig 4b)** and identified that for scenarios when VE_DIS_ =90% is mediated mostly by VE_SYMP_ (90%, black circles), increasing VE_INF_ from 10% to 90% resulted in substantial further reductions in diagnosed cases (∼20,000) and deaths (∼60). When VE_DIS_=90% was mediated entirely (90%, white circles) or mostly (70%, grey circles) by VE_SUSC_, then increasing VE_INF_ from 10% to 90% resulted in very limited further reductions in diagnosed cases and or deaths.

**Figure 4.**
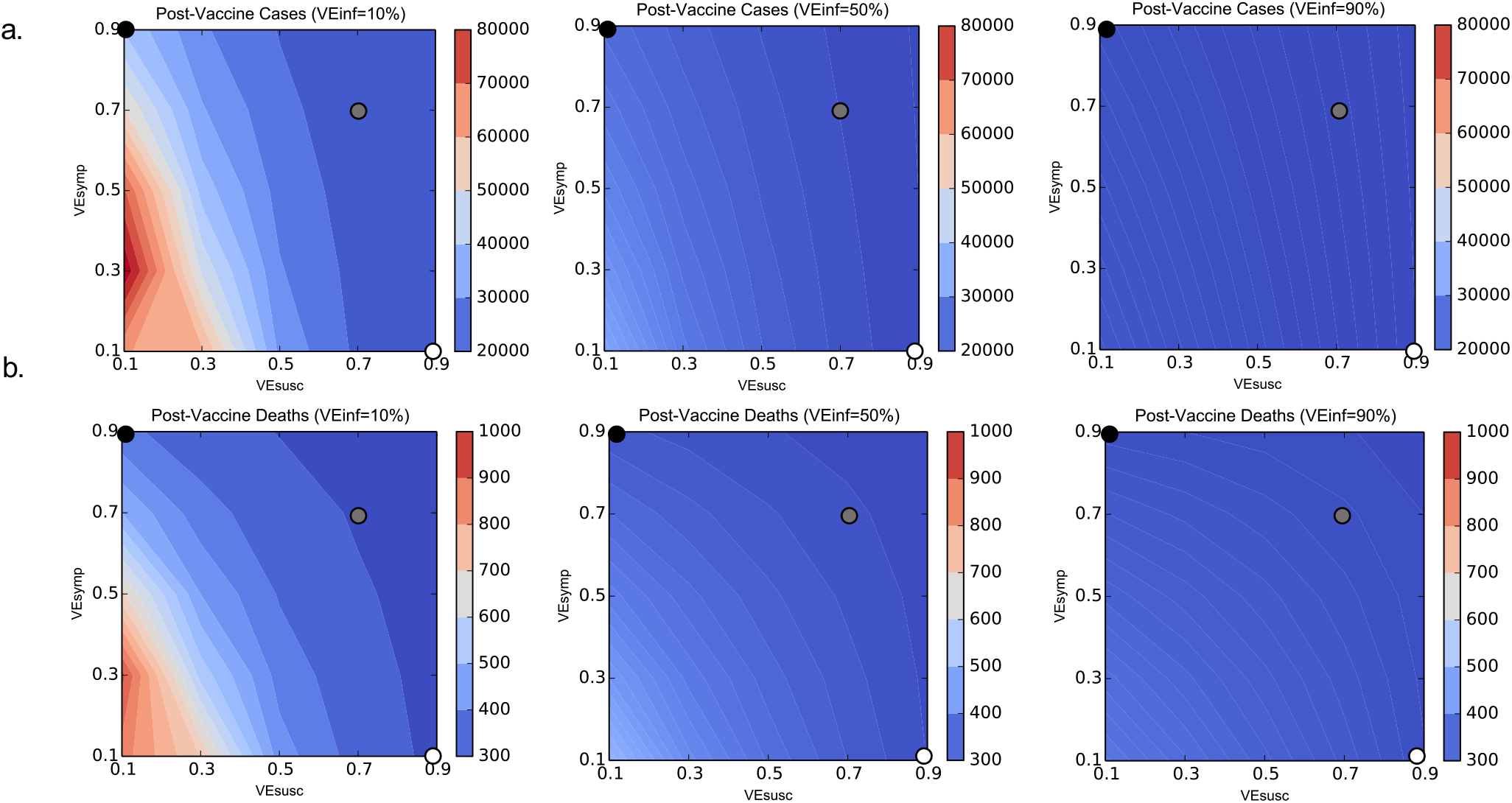
High VE_INF_ further reduces infections and death only when VE_SUSC_ is low or moderate. Heat maps comparing contrasting vaccine scenarios. **a**. Post-vaccine diagnosed cases (top row) and **b**. post-vaccine deaths (bottom row) with different combinations of VE_SUSC_ and VE_SYMP_. In this simulation, there were 62979 diagnosed cases and 1144 deaths prior to vaccination and heat maps capture all outcomes beyond this point. The left column assumes VE_INF_=10%; middle column assumes VE_INF_=50%; right column assumes VE_INF_=90%. The dots are 3 scenarios compatible with results from the Pfizer and Moderna trials in which VE_DIS_=90% (black is VE_SYMP_ = 90% / VE_SUSC_ = 0%, grey is VE_SYMP_ = 70% / VE_SUSC_ = 70% and white is VE_SYMP_ = 0% / VE_SUSC_ = 90%). Increased VE_INF_ leads to a substantial further reduction in cases when VE_DIS_ is mediated by high VE_SYMP_ but not when it is mediated by high VE_SUSC_. In general, additional benefit of VE_INF_ is accrued when VE_SUSC_ is low, across a wide range of VE_SYMP_.

We next identified that VE_SUSC_ and VE_INF_ had nearly equivalent effects on number of post-vaccine diagnosed cases **(Sup fig 4a)** and deaths **(Sup fig 4b)**. For VE_SYMP_ = 10% and 50% (**Sup fig 4, left and middle columns**), at values of VE_SUSC_<50%, increases in VE_INF_ lead to further reductions in cases and deaths. However, at high values of VE_SUSC_, increases in VE_INF_ added little benefit. There was moderate effect modification by VE_SYMP_: particularly for deaths, there was less added benefit of increasing VE_INF_, when VE_SYMP_ = 90% (**Sup fig 4, right column**) highlighting that VE_SYMP_ prevents deaths more efficiently than infections.

### VE_INF_ as a determinant of the severity of a fourth wave in the event of slow vaccine roll out

The distribution and acceptability of vaccines to the public remains uncertain. We therefore simulated the vaccine scenarios in **Fig 2** assuming half (2500 vaccines / day with completion in March 2022, **Fig 5, left column)** the roll out speed. Here we assumed VE_SYMP_=90% such that all nine considered scenarios had efficacies compatible with the Moderna and Pfizer results. Nevertheless, at slower roll out, a fourth wave of infections **(Fig 5a)**, diagnosed cases **(Fig 5b)**, hospitalization **(Fig 5c)** and deaths **(Fig 5b)** occurred among all scenarios. The peak **(Fig 5 top row)** was blunted and delayed under scenarios with VE_SUSC_=90% (blue lines) or VE_INF_=90% (dark red, green and blue) with ∼100,000 fewer cumulative cases and ∼300 fewer deaths **(Fig 5 middle and bottom rows)** indicating that VE_INF_ would take on added importance under less optimal roll out scenarios with low VE_SUSC_. Only the scenario with limited effects on secondary transmission VE_SUSC_=10% and VE_INF_=10% allowed a severe enough wave to necessitate another round of physical distancing **(Fig 5e,f)**.

**Figure 5.**
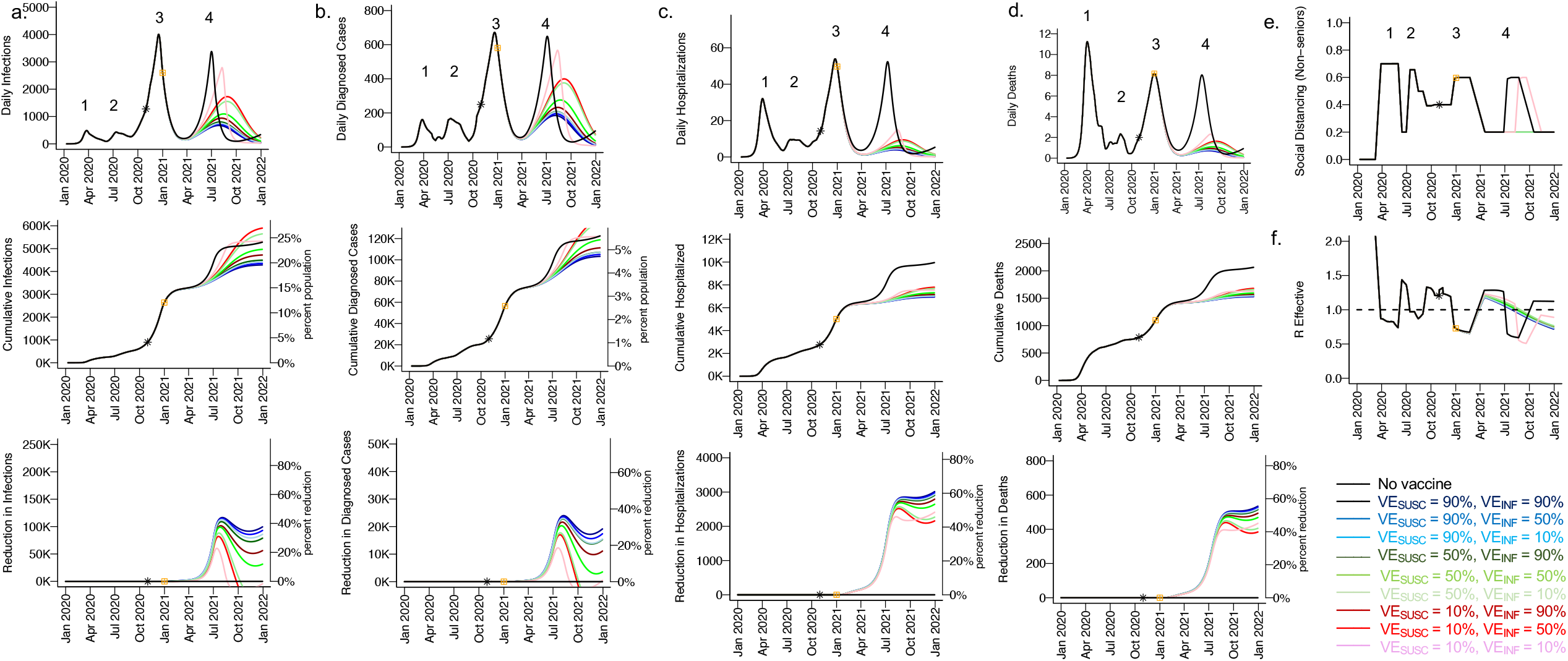
High VE_SUSC_ or high VE_INF_ limit cases and deaths at low vaccine roll out rates. For unvaccinated (black lines) and each vaccine cohort (colored lines, legend), we project **a**. infections, **b**. diagnosed cases, **c**. hospitalizations and **d**. deaths, as well as **e**. social distancing relative to pre-pandemic levels and **f**. the effective reproductive number. The first four columns are organized by row: top = daily incidence, middle = cumulative, bottom = reduction since day of vaccination. Waves of infection are numbered 1-4. Nine combinations of VE_SUSC_ and _F_ are considered while VE_SYMP_ is fixed at 90% such that all vaccines would produce results consistent with those in the Pfizer and Moderna trials. High VE_SUSC_ (90%) simulations are and have similar outcomes to one another. Moderate VE_SUSC_ (50%) simulations are green. Low VE_SUSC_ (10%) simulations are red / pink. Dark lines are high VE_INF_ (90%) and have similar outcomes to one another. Moderate darkness lines are medium VE_INF_ (50%). Light lines are low VE_INF_ (10%). The largest reduction in cases is associated with either high VE_SUSC_ or _F_. 2500 vaccines are given per day starting January 1, 2021 (yellow square) until 50% are vaccinated.. Case threshold for reinstituting physical distancing to 0.6 is 300 per 100,000 and relaxation is 100 per 100,000. 80% of vaccines are initially allocated to the elderly with the remaining 20% to middle-aged cohorts.

Vaccine uptake at 5000 per day resulted in R_eff_<1 in April to June at 20% social distancing relative to pre-pandemic conditions (SD=0.2), a rough proxy for herd immunity assuming some degree of residual social distancing and masking **(Fig 6a)**. Vaccine uptake at 2500 per day achieved R_eff_<1 in August to September **(Fig 6b)**. The more rapid roll out scenario achieved this functional herd immunity before occurrence of the fourth wave whereas under the slower roll out scenario, a surge in cases in July contributed to development of R_eff_<1 (**Fig 6, bottom row)**. Under both roll out rate scenarios, vaccines that had both low VE_SUSC_ and VE_INF_ (light green and pink), had a one-to-two month delay to reach R_eff_<1, thereby allowing more infections.

**Figure 6.**
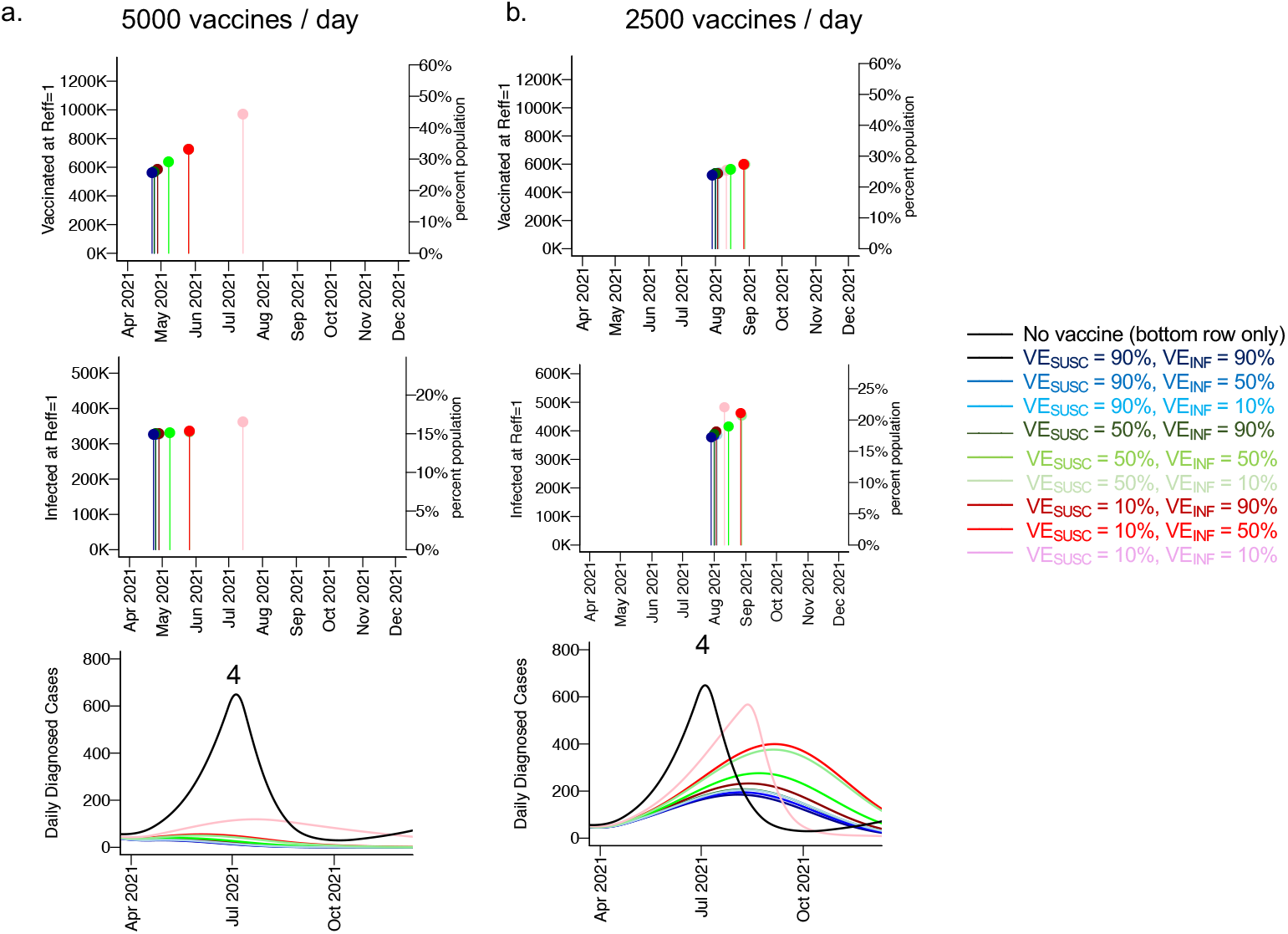
Slow vaccine rollout rate and a vaccine with low VE_SUSC_ and VE_INF_ would prolong time to functional herd immunity. **a**. 5000 vaccines are given per day until 50% are vaccinated. **b**. 2500 vaccines are given per day until 50% are vaccinated. The top row is number of vaccinated people (y-axis) and date (x-axis) at which R_e_ decreases below 1. Middle row is cumulative number of infected people (y-axis) and date (x-axis) at which R_e_ decreases below 1. Bottom row is the projected 4^th^ wave at these scenarios demonstrating that ongoing cases contribute to herd immunity with slower roll out only. VE_SYMP_=90% is assumed for each of the nine vaccines. The most rapid time to achieve herd immunity is associated with either high VE_SUSC_ or VE_INF_. Of note, these simulations occur at 20% social distancing.

### VE_INF_ as a determinant of whether a fourth wave of cases occurs in the event of waning vaccine efficacy

The duration of SARS-CoV-2 vaccine induced immunity also remains uncertain. We simulated the vaccine scenarios in **Fig 2** but with VE_SYMP_=90% assuming slow (1% per month, **Sup fig 5, left column)**, moderate (5% per month, **Sup fig 5, middle column)**, and rapid (10% per month, **Sup fig 5, right column)** reversion from vaccine immune to susceptible. At high loss of efficacy, a fourth wave of cases **(Sup fig 5a)** and deaths **(Sup fig 5b)** eventually occurred among all vaccine scenarios, but was delayed under scenarios with VE_SUSC_=90% (blue lines) or VE_INF_=90% (dark red, green and blue) indicating that VE_INF_ may also take on critical importance in the event of waning vaccine efficacy.

### A virologic mathematical model of SARS-CoV-2 transmission

The above results suggest that the potential severity of a fourth wave can only be estimated with accurate estimates for VE_SUSC_, VE_SYMP_ and VE_INF_ among relevant vaccines, as well as rates of vaccine roll out and duration of protection. It is therefore a priority to identify the true values for these vaccine characteristics.

Based on experience from multiple other viruses that exposure dose predicts transmission (*30, 31*), we hypothesize that VE_INF_ is likely to be mediated by reduction in viral load among infected people **(Fig 7)**. We therefore employed our existing intra-host model described in the **Methods** that links SARS-CoV-2 viral load dynamics in an infected person with the potential for transmission. The model reproduces two key distributions: the heterogeneous number of people secondarily infected by each person (termed *individual R0*) and the serial intervals associated with each transmission pair (*16, 32, 33*). The resulting model output explains observed super-spreader phenomenon, and predicts a transmission dose response curve which captures the probability of transmission given an exposure viral load (*14, 26*). The model was compatible with existing data showing that most transmissions occur during the pre-symptomatic phase of infection (*16*).

**Figure 7.**
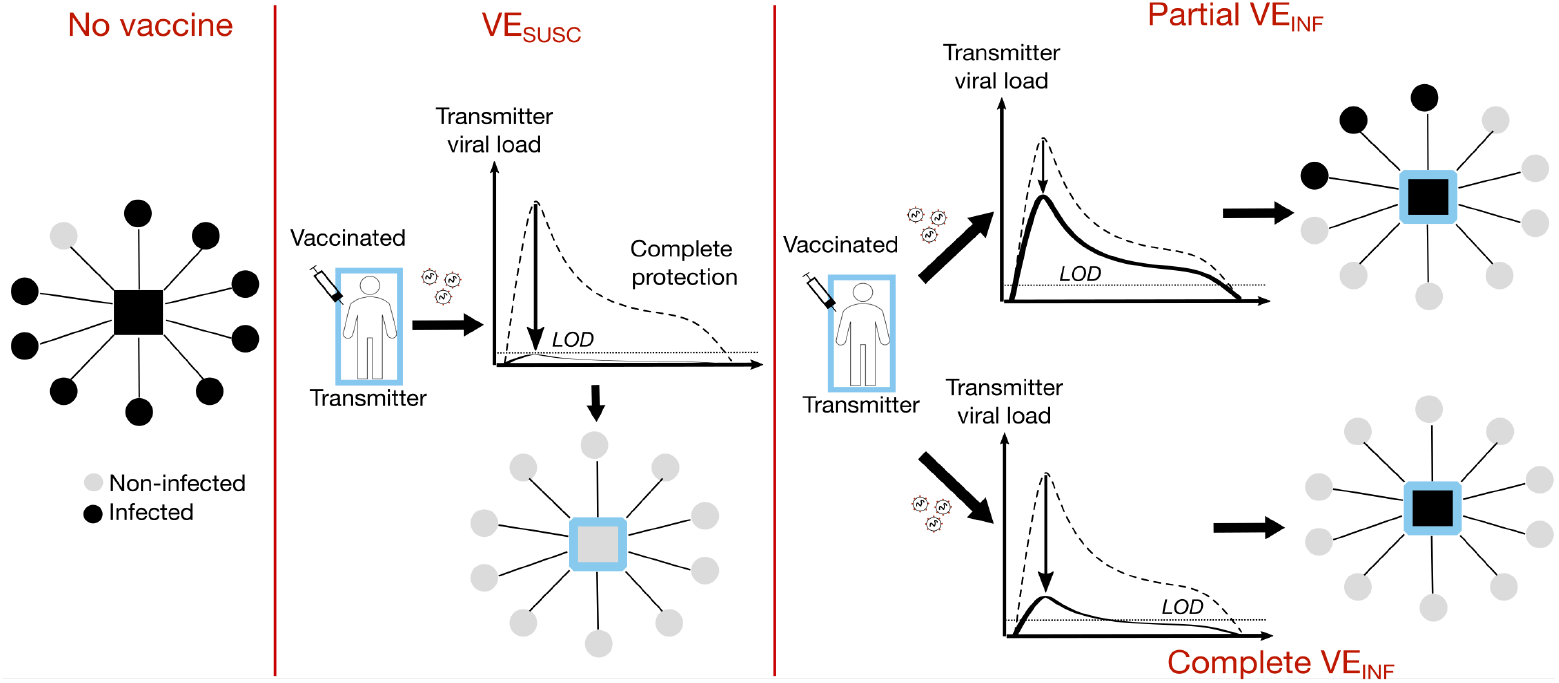
Conceptual basis for reduction in viral load lowering transmission.

### Small reduction in peak viral load required for lowering VE_INF_

We next considered methods to estimate VE_INF_ using viral load as a potential surrogate. To simulate the effect of a vaccine on viral load, we assumed the presence of a tissue-resident population of immune cells with rapid ability to recognize and kill infected cells, as well as proliferate *in situ*, as a necessary condition to lower peak viral load in infections such as SARS-CoV-2 with rapid initial growth kinetics (*34*). We first established a relationship between the initial number of tissue resident immune cells and peak viral load **(Fig 8a, b)** during individual simulated infections. We then assumed vaccination trials consisting of 1000 people in which vaccine recipients generated a certain number of these immune cells while placebo recipients did not. By estimating the reduction in number of transmissions, we then were able to estimate VE_INF_ for each vaccine. The model predicted a sigmoidal relationship between reduction in peak viral load and VE_INF_: a 0.6 log or 4-fold reduction in peak viral load resulted in VE_INF_ =50% and a 2.5 log or ∼300-fold reduction resulted in VE_INF_ = 90% **(Fig 8c)**.

**Figure 8.**
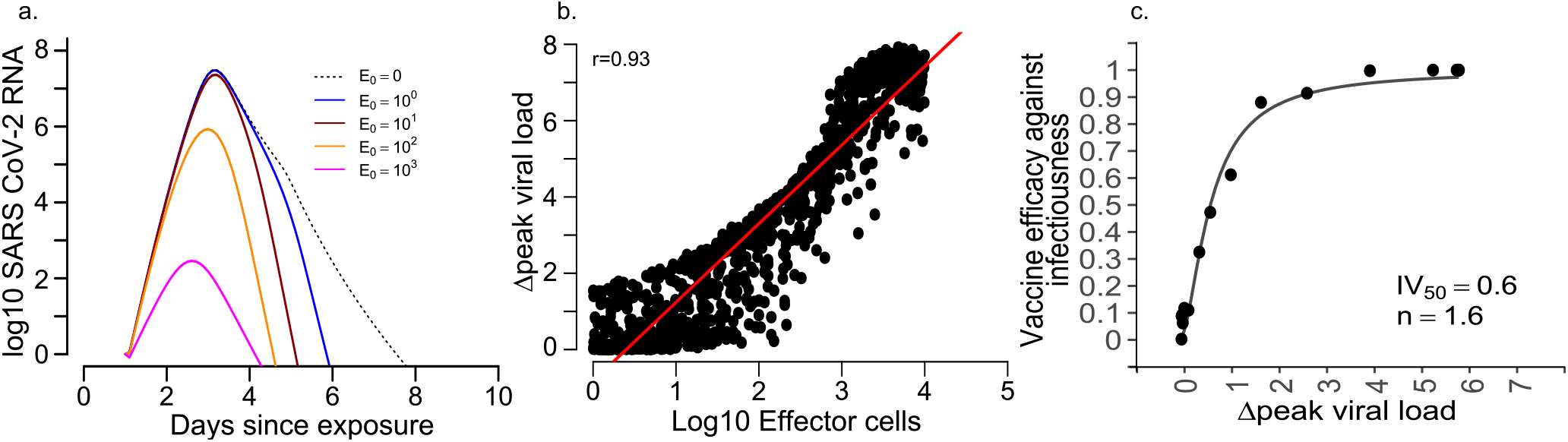
Small reduction in peak viral load due to vaccinations would translate to significant VE_INF_. **a**. Simulated virologic trajectories with higher imputed initial number of vaccine-generated tissue-resident immune cells (E_0_) demonstrate lower peak viral loads. **b**. Varying number of tissue-resident immune effector cells generated by a vaccine (x-axis) predicts peak viral load (y-axis) in individual infection simulations each denoted with a dot. The red line indicates a correlation line. **c**. Reduction in viral load (x-axis) predicts VE_INF_ (y-axis) in vaccine simulations. Each black dot is a simulation of 1000 vaccine recipients given a vaccine which generates a fixed E0 versus 1000 placebo recipients. VE_INF_=50% is achieved with a 0.6 log_10_ reduction in peak viral load. VE_INF_=90% is achieved with a 2.5 log_10_ reduction in peak viral load. The relationship between change in peak viral load (x) and VE_INF_ is captured with the formula: VE_INF_ = (log_10_ x)^1.6^ / (IV50)^1.6^ + (log_10_ x)^1.6^ where IV50=0.6.

### Proposed animal challenge experiments for establishing transmission dose required for SARS-CoV-2

Our model derived prediction that vaccine induced reduction in exposure viral load could serve as a surrogate endpoint for VE_INF_ requires experimental validation. While a transmission dose response relationship has been demonstrated for SARS-CoV-1 in mice (*35*), and infection of SARS-CoV-2 between golden hamsters occurred with direct viral load exposure at 10^11^ RNA copies but not 10^9^ viral RNA copies (*36, 37*), formal dose response experiments are still needed for SARS-CoV-2. We performed mock experiments in which 30 animals (either mice, hamsters or non-human primates) are challenged with 3 or 5 graded doses of viruses selected from experience with SARS-CoV-1. This method demonstrated that given the existence of a true transmission dose response curve, we can identify the slope and ID50 (dose at which 50% transmission occurs) >99% of the time if 5 doses (selected based on the TD50 and slope from SARS-CoV-1 challenge in mice) are used in the experiments **(Sup table 5)**. If the initial estimate for ID50 is mis-specified 20-fold, then the failure percentage increases to 7% and the 95% confidence range for slope and ID50 of the curve are wider **(Sup table 6)**. However, if we employ a dynamic sample size allocation procedure where we determine the challenge dose based on the number of infections in a single first dose group (subsequent doses are increased if all mice remain uninfected or vice versa) then the uncertainty range for both parameters decreases and the failure percentage is miniscule **(Sup table 7)**. Overall, this result suggests that the transmission dose response curve predicted by our mathematical model could be verified quickly in graded challenge experiments in animals, adding further veracity to viral load as a reasonable proxy for transmission risk in clinical trials.

### Proposed study design for randomized placebo controlled double blinded human challenge vaccines trials with viral load as endpoints

One method to directly assess VE_SUSC_ and VE_SYMP,_ and to indirectly estimate VE_INF_, using projections from our model **(Fig 8c)** is a human challenge study in which healthy participants are challenged with SARS-CoV-2 one month after receiving a final dose of a vaccine or placebo schedule in a blinded fashion. Following challenge, viral load would be sampled daily for 2 weeks to capture true peak.

Under this study design, we would expect all placebo recipients to develop virologically confirmed infection and for most to be symptomatic. Assuming VE_DIS_=90%, then only ∼10% of vaccine recipients would develop symptoms with a confirmatory PCR test. If VE_DIS_ is mediated entirely by VE_SUSC_, then vaccine recipients would shed no virus as in **Fig 7** and the study would not be able to assess VE_INF_. However, as shown conceptually in **Fig 1** and in projected model data in **Figs 4**, VE_INF_ has little added benefit when VE_SUSC_ is high making its estimation unnecessary for population projections.

If VE_DIS_ is mediated entirely by VE_SYMP_, then vaccine recipients will shed virus as in **Fig 7** and useful comparisons can then be made between infected vaccine and placebo recipients. We demonstrate that with this approach, 80% power can be achieved to detect differences in peak log_10_ viral load between vaccine and placebo arms with as few as 10 infected participants per arm if peak viral reduction is 2.5 log_10_ (model estimated VE_INF_∼90%), 52 participants per arm if peak viral reduction is 1.0 log_10_ (model estimated VE_INF_∼60%), and 143 participants per arm if peak viral reduction is 0.6 log_10_ (model estimated VE_INF_=50%) **(Sup fig 6)**.

## Discussion

An optimal vaccine program would prevent the maximum numbers of cases and deaths, without the need for further lockdown periods. The first component of such a program will be testing and licensing of vaccines that provide at least partial protection from symptomatic disease (VE_DIS_). Initial data from the Pfizer and Moderna trials suggest that these products may have 90% or higher VE_DIS_. The second step is to consider the proportion of the population that will need to be vaccinated in order to provide herd immunity. This threshold will depend critically on indirect effects that protect unvaccinated members of the population. Indirect effects occur when VE_DIS_ is mediated by VE_SUSC_ rather than VE_SYMP_, but may also be augmented by a vaccine product with high VE_INF_ in the scenario where VE_SUSC_ is low. Given rapid enough roll out, our results suggest that vaccines with either high VE_SUSC_ or high VE_INF,_ or both moderate VE_SUSC_ and moderate VE_INF_, could prevent a fourth wave of cases and deaths in the spring and summer.

Unfortunately, VE_SUSC_ can only be partially discriminated from VE_SYMP_ in current clinical trials using serologic assays which may miss infection due to waning humoral responses (*38*). Moreover, VE_INF_ is not being directly assessed. While viral load is being compared between symptomatic infected vaccine and placebo recipients, in most cases, sampling will occur several days after the peak viral load when transmission risk is highest (*1*). Viral load in asymptomatic cases also will not be captured. Overall, VE_SUSC_, VE_SYMP_ and VE_INF_ are particularly challenging to measure, leaving policy makers with incomplete information for projecting the impact of a given vaccine.

We identify that under any scenario in which VE_SUSC_ is low, a vaccine with VE_INF_ >50% would add substantial protection at the population level. A vaccine with this profile would exert maximal benefit whether given to younger or elderly cohorts first if rolled out quickly enough. If VE_SYMP_ is driving observed results, then high VE_INF_ would be vital for preventing thousands of cases and saving hundreds of lives in King County, with much larger benefits when considered across the larger US population. In scenarios where a fourth spring wave is inevitable, such as slow vaccine roll-out or waning vaccine induced protection, VE_INF_ could potentially delay and blunt the peak number of cases and deaths, thereby preventing the need for re-enforcement of physical distancing while also preventing many deaths.

Therefore, it is an urgent research priority to identify reasonable estimates for VE_SUSC_, VE_SYMP_ and VE_INF_ for vaccines that will be given to the population at large. Trial strategies which attempt to directly measure secondary infections in households (*39, 40*), or to assess the degree of protection afforded to unvaccinated members of communities with partial vaccination relative to communities with less vaccination, would potentially be conclusive. They will need to be performed quickly to obtain actionable results prior to spring 2021.

Our analyses suggest that peak viral load could serve as a surrogate endpoint for secondary transmission and allow for rapid, complementary studies. We estimated the relationship between viral load and transmission probability for SARS-CoV-2 based on model fitting to observed serial intervals and individual R0 values (*14*). The emergent transmission response curve took on a similar sigmoidal shape to empirically derived curves for SARS-CoV-1 in a controlled set of murine experiments (*35*), and also resembled the relationship between quantitative viral PCR and probability of culture positivity in humans infected with SARS-CoV-2 (*41*).

As a first step, it is necessary to formally test the hypothesis that exposure viral load is predictive of transmission risk (*42*). A valid viral load surrogate cannot currently be inferred from human cohorts as the exposure viral load is never documented between transmission pairs, though formal surrogate endpoint analysis will ultimately be necessary if sufficient data emerges. We suggest that animal models of infection are ideal for this purpose and that necessary transmission dose can be inferred with a relatively small number of non-human primates or mice.

Human trials using reduction in peak viral load or viral area under the curve as correlates for reduction in VE_INF_ could take one of two forms. The first would involve prospective nasal sampling of virus in all enrolled participants with virologic endpoints compared between those who become infected in vaccine and placebo arms. An ideal study population would be university students due to their high incidence rate and low overall infection morbidity. The advantages of this approach would be real-world validation of biologic vaccine effects in which participants experience natural variability in potentially critical factors such as viral exposure dose, time between vaccination and infection, and route of transmission. The relationship between viral load and symptoms would also be clarified with this study design. Challenges would be operational including large samples size and a massive number of prospective samples.

Human challenge studies are a potentially rapid method to directly measure VE_SUSC_ and VE_SYMP,_ and to indirectly estimate VE_INF_ using viral load, as each participant would contribute to the study endpoints. This approach could potentially be completed in at minimum 104 participants within 2-3 months, depending on the selected time between vaccination and viral challenge. While challenge studies are most efficient, there are important ethical considerations regarding potential harm to study participants. Moreover, it will be uncertain whether results can be generalized to the wider population, particularly those in different age cohorts. Nevertheless, even crude estimates of VE_SUSC_, VE_SYMP_ and VE_INF_ could add critical knowledge to influence vaccine implementation policies.

Our approach has limitations. We combine several scales of models which reflect population conditions unique to King County Washington and virologic findings from across the globe. The models are not equipped to make precise vaccine schedule assessments for different locations and are not meant as predictions. Rather, we intend to make the conclusion that VE_INF_ could theoretically provide substantial population level benefits and to provide a framework for most rapid evaluation of this metric. The scope of the ongoing third wave is difficult to forecast and will depend on changes in human behavior over the next several weeks. The number of cases and deaths during a possible fourth spring wave may be somewhat dependent on current events.

In conclusion, in the situation where observed high VE_DIS_ is predominately due to reduction in symptoms rather than absolute protection against infection, VE_INF_ will be vital to measure as it may determine whether a severe fourth wave of cases and deaths is imminent in the spring. Using peak viral load as a proxy measure in human challenge studies is an efficient way to complement other clinical trial designs to assess VE_INF_.

## Methods

### King Country transmission model

We modified a previously developed deterministic compartment model (*43*), which captures the epidemic dynamics in King County, WA between January 2020 and October 2020 and projects the trajectory of the local pandemic through the end of 2021 in the absence and presence of vaccines. Vaccination is simulated with a starting date of January 1, 2021. Our model stratifies the population by age (0-19 years, 20-49 years, 50-69 years, and 70+ years), infection status (susceptible, exposed, asymptomatic, pre-symptomatic, symptomatic, recovered), clinical status (undiagnosed, diagnosed, hospitalized) and vaccination status.

In our main scenario we assume that 20% of infections are asymptomatic and that asymptomatic people are as infectious as symptomatic individuals but missing the highly infectious pre-symptomatic phase. As a result, the relative infectiousness of individuals who never develop symptoms is 56% of the overall infectiousness of individuals who develop symptomatic COVID-19. This conservative estimate falls between the 35% relative infectiousness estimated in recent review based on 79 studies (*44*) and the current best estimate of 75% suggested by the CDC in their COVID-19 pandemic planning scenarios (*45*).

The forces of infection, representing the risk of the susceptible individuals to acquire infection (transition from susceptible to exposed), are differentiated by age of the susceptible individual, the contact matrix (proportion of contacts with each age group), infection and treatment status (asymptomatic, pre-symptomatic, symptomatic, diagnosed and hospitalized cases) of the infected contacts as described in the **Supplement**.

The model is parameterized with local demographic and contact data from King County, WA and calibrated to local case and mortality data using transmission parameters ranges informed from published sources (*15, 46-48*).

A critical parameter in the model is the social distancing metric which estimates the amount of potential infection contacts between members of the population. This parameter is intended to capture physical contact reduction due to physical distancing policies, but also decreased number of transmission contacts due to masking. The parameter varies between 0, which represent pre-pandemic level of interactivity, and 1, which represents complete physical distancing with no interactivity.

In the current version, we incorporate fluctuating values of this parameter retrospectively to calibrate the model to observed infection, hospitalization and death data through the end of October 2021, as well as prospectively to capture likely enhanced physical distancing in response to present and future cases. Our benchmarks for increasing physical distancing to 0.6 was when 2-week average number of cases exceeded 300 per 100,000. We allowed relaxation of the parameter to 0.2 when 2-week average number of cases fell below 100 per 100,000. For elderly populations, we assume greater restrictions to 0.8 and lessen relaxation to 0.4. This approach reproduces the waves of infection which have defined the United States pandemic to date.

### Vaccine simulations in King County, Washington

We consider several vaccine efficacy profiles as described in the **Results** with different efficacies as defined in **Table 1**. Implementation of these efficacies is described in the **Supplement**.

### SARS-CoV-2 within-host model

We next employed a within-host model describing SARS-CoV-2 infection from our previous study to generate viral loads to assess transmission risk (*21*). The viral load generating model is included in the **Supplement**.

### Intra-host transmission model

We employed our previously described model linking transmitter viral load with probability of transmission (*21*). The details of this model are described in the **Supplement**.

### Intra-host model vaccination simulations

We simulated the impact of the vaccination by assuming that a vaccine generates a certain number of SARS-CoV-2 specific acquired immune cells that are ready to proliferate (with no need for precursor compartments *M*_1_ and *M*_2_) and act to quickly eliminate the ongoing infection. We thereby modify our intra-host model in the **Supplement** as,

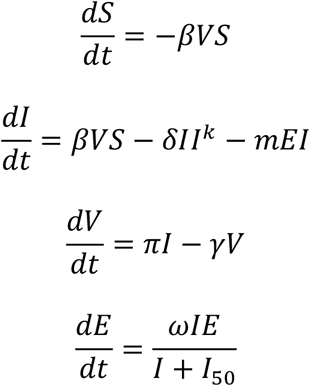

Here, *I*_50_ denotes the level of infected cell that allows proliferation of immune cells at 50% maximal. We assume it to be 10 cells/mL. We further fix ω = 2 days^-1^cells^-1^ and *m* = 0.01 days^-1^cells^-1^.

Each vaccine trial consists of selecting a starting condition of parameter E (E_0_) that leads to a predictable reduction in peak viral load **(Fig 6b)**. We simulate individual trials with 1000 vaccine participants and 1000 placebo recipients, and then assess the relative reduction in transmissions to estimate VE_INF_ as in **Table 1**.

### Graded challenge sample size estimates

We assume that the dose response model is given by 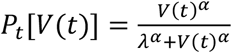 where *P_t_*[*V*(*t*)] is the infectiousness based on viral loads V (dose) at the time t of viral challenge, λ is the infectivity parameter that represents the viral load that corresponds to 50% infectiousness and α is the Hill coefficient that controls the sharpness in the dose-response curve.

We consider the trial design given this initial estimation of parameters: α = 0.8 (from our human model and λ = 100 pfu from murine experiments with SARS-CoV-1 (*35*). We estimate α and λ under the given total sample size (N = 30) with different combination of dose assignment. Particularly, we consider five does: V_1_ = ID10 = 6, V_2_ = ID30 = 35, V_3_ = ID50 = 100, V_4_ = ID70 = 290, and V_5_ = ID90 = 1600 pfu. We consider three different settings to allocate sample size: (1) equal sample size among the five dose (6 animals each dose group); (2) equal sample size among V_1_, V_3_, and V_5_, (10 animals each dose group; and (3) equal sample size among V_2_, V_3_, and V_4_. We simulate the data from the two-parameter dose-response model and use the function ‘drm’ in the R package ‘drc’ to fit the model.

We consider sensitivity analysis when ID50 is poorly specified, especially if ID50 is smaller than expected. We also we consider a dynamic sample size allocation procedure. Particularly, we first allocate n1 = N/5 animals to V_1_ and determine the following dose allocation based on the number of infections in this V1 dose group, denoted as m1. If more than half of the animals allocated to V_1_ are infected, i.e., m1 ≥ n1/2, we set λ∗ = V1 and assign the rest of the animals equally to the doses V_2_^∗^ =1pfu, V_3_^∗^ =2pfu, V_4_^∗^ =18pfu and V_5_^∗^ =100pfu, which reflect this new λ^∗^. Based on this dynamic sample size allocation procedure, the proportion of replicates that fail to produce an estimate is smaller and we are able to identify the values of α and ID50 with increased accuracy.

### Human challenge study sample size estimates

Sample size calculations were based on a 2-sample t-test to compare mean peak log_10_ viral load between the vaccine and placebo arm with 80% power. We conservatively assumed a common standard deviation of 1.8 log_10_ for the peak log_10_ viral load in both arms, which is based on estimates from natural infection (*49*), and likely an upper limit for our proposed human challenge trial where the challenge dose and anatomic site would be equivalent. We also assumed equal number of participants in each arm, and a 2-sided type I error of 0.05. We considered a range of values for the difference between peak log_10_ viral loads in the vaccine and placebo arm that corresponded to projected values of VE_INF_ ranging from 30% to 100%.

## Data Availability

Code for our mathematical model will be provided immediately upon request.

## Supplementary materials

### King County Mathematical Model of SARS-CoV-2

Our mathematical model consists of a series of differential equations (*1*), which describe a series of compartments described in **Sup fig 1** and in the **Results**. Equations are listed here:

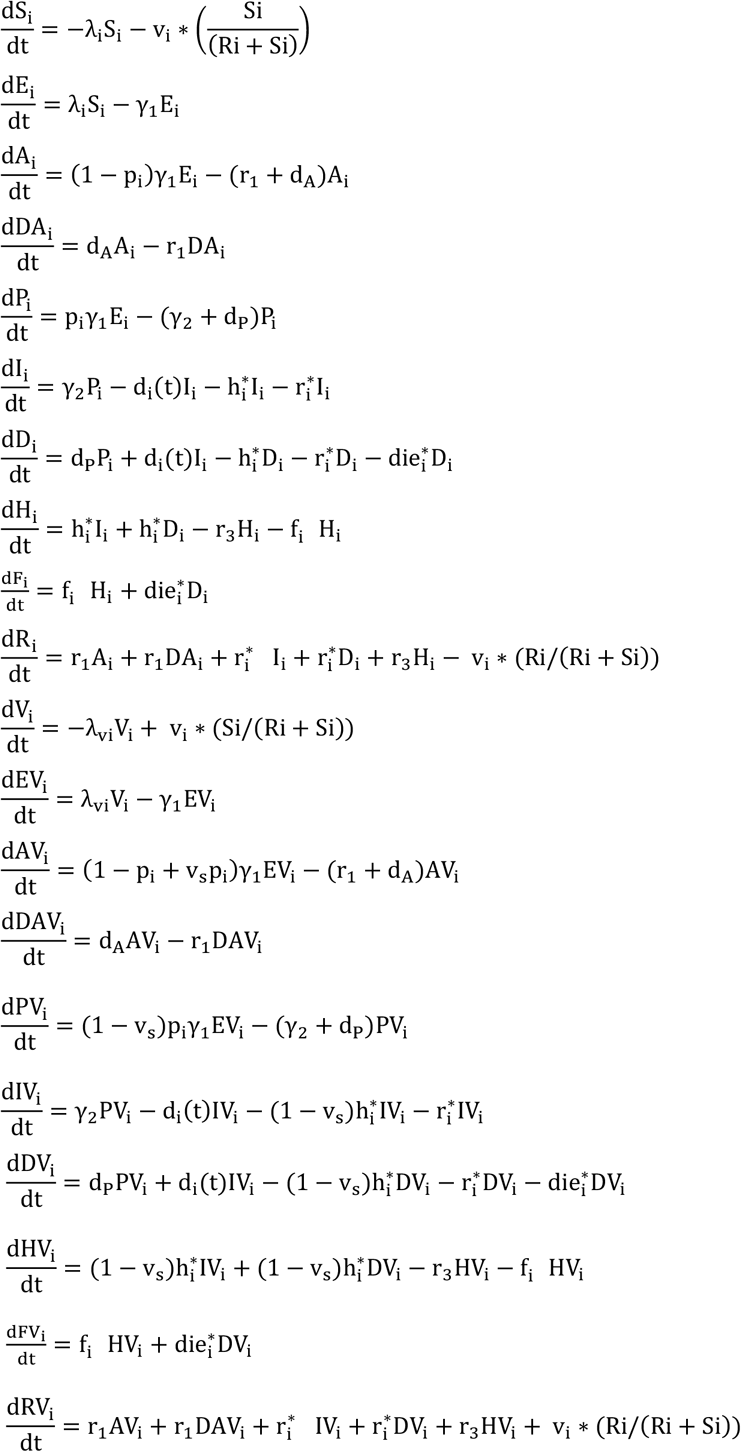

### King County model parameters

Model parameters are listed as follows with values listed in the **Supplementary Table 1:**

p_i_ – proportion of the infections which become symptomatic by age in absence of a vaccine γ_1,_ γ_2_-progression rates from exposed (E) to infectious (A and P) to symptomatic (I)

h_i_ – hospitalization rate among severe cases by age in absence of a vaccine

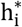 – hospitalization rate among diagnosed by age (calculated)

r_1-3_ - recovery rate of the asymptomatic, mild symptomatic and hospitalized cases

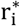 - recovery rate of the diagnosed symptomatic cases by age (calculated)

f_i_ – fatality rate among hospitalized by age

v_i_ – vaccination rate by age including prioritization strategy

v_s_ – vaccine efficacy in reducing the risk of symptomatic infection upon acquisition

v_h_ – vaccine efficacy in reducing the risk of hospitalization upon symptomatic infection

die_i_ – death rate without hospitalization by age.

d_i_ – diagnostic rate by age.

The diagnostic rates are estimated using testing data from the Washington State Department of Health (DOH). The equation uses the average daily tests for the time period in question divided by an estimate of the number of people desiring tests. The rates are divided across age groups according to the fraction of tests in each age group during that month. To get the test demand estimate, we fit a parameter rho_S that represents to likelihood of non-infected individual seeking a test compared to a symptomatically infected individual.

The forces of infection (λ_i_), representing the risk of the susceptible individuals by age to acquire infection (transition from susceptible to exposed), are differentiated by age of the susceptible individual, a contact matrix (proportion of contacts with each age group), infection and treatment status (asymptomatic, pre-symptomatic, symptomatic, diagnosed and hospitalized cases) of the infected contacts, and the time-dependent reduction of transmission due to physical distancing measures (work from home, closing non-essential businesses, banning large gathering, etc.) applied in the area (scaled up starting March 8 and fully taking effect March 29) and later relaxed during the reopening after May 15 to values that are fit monthly until Oct 31^st^, 2020.

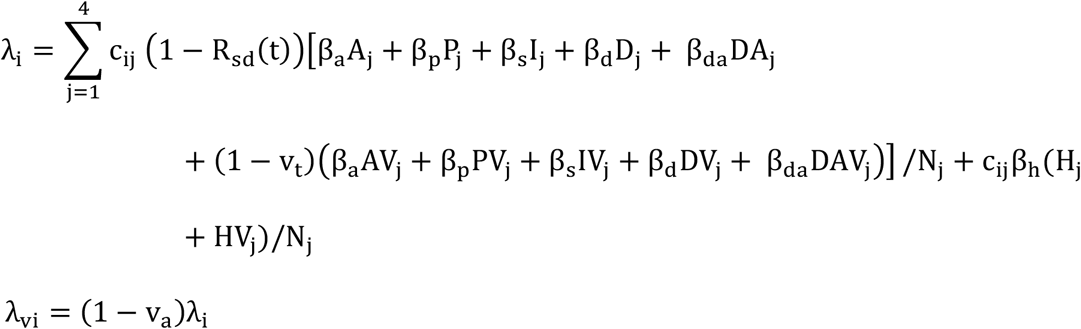

where β_a_, β_p_, β_s_, β_d_, β_h_ are the transmission rates from contacts with asymptomatic, pre-symptomatic, symptomatic, diagnosed and hospitalized infections (before the start of COVID measures at t= δ_1_), c_ij_ is contact matrix (proportion of the contact with other age groups), N_i_ is population size by age, v_a_ is vaccine efficacy in reducing the acquisition risk (reduction of susceptibility), v_t_ is vaccine efficacy in reducing the transmission risk (reduction of infectiousness).

R_sd_ (t) is the reduction of transmission due to physical distancing and other preventive measures which is applied uniformly to all age groups. It is scaled up linearly from 0 to 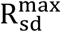 between t= δ_1_ and t= δ_2_). Later it is calibrated monthly to match the King County epidemic through October of 2020 and then controlled dynamically based on the bi-weekly case rates per 100k of the population. For age groups 1-3 the highest value of 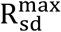 is 0.6 (i.e. interactions at 40% of pre-COVID levels) and the lowest allowed is 0.2 (social interactions at 80% of pre-COVID levels). These limits are each 0.2 higher for the oldest age group. The triggers for increasing or decreasing social distancing levels are given in the parameter table.

### King County model calibration

The model is calibrated to 3 “targets” based on local data (**Supplementary fig 2**), namely: the age-wise number of confirmed daily cases (**S2d**), daily deaths (**S2e**) and daily hospital admissions (**S2f**) reported in King County over time since the start of the epidemic outbreak through October 31^st^, 2020. We used the BFGS optimization algorithm to estimate the best parameter values for the time period being fitted. We defined thresholds for each parameter and proceeded with the best set reported by the routine. Calibration was divided into multiple periods. The first was from the start of the epidemic through the initial lockdown period ending in early May. Subsequent fits were by month, but sharde the initial fits for start date, β_∗_ (overall infectivity) and β_d_ (adjustment to infectivity for diagnosed individuals).

### Intra-host model of SARS-CoV-2 kinetics

This model assumes SARS-CoV-2 (*V*) infects susceptible cells (*S*) at rate *β* producing infected cells (*I*) that then generate new virus at a per-capita rate *π*. In the model, the death of infected cells is mediated by (1) the innate responses *δI*^*k*^ which is dependent on the infected cell density and the exponent *k*, (2) the acquired immune responses 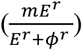 by SARS-CoV-2-specific effector cells (*E*). The acquired responses are non-linear and are captured by the Hill coefficient *r* that allows rapid saturation of the killing of infected cells whereas the parameter *ϕ* determines the level of SARS-CoV-2-specific effector cells at which the saturation occurs. Moreover, we describe the rise of SARS-CoV-2-specific effector cells in a two-stage manner. The first stage defines the proliferation of the first precursor cell compartment (*M*_1_) at rate *ω* and differentiation into a second precursor cell compartment (*M*_2_) at a per capita rate *q*. Finally, second precursor cells differentiate into effector cells at the same per capita rate *q* and are cleared at rate *δ*_*E*:_. More details on the model can be found in the ref (*2*).

The model is expressed as a system of ordinary differential equations:

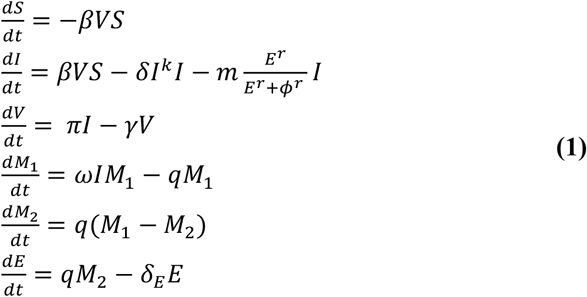

The initial conditions for the model were assumed as *S*(0) = 10^7^ cells/mL, *I*(0) = 1 cells/mL, 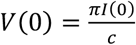, *M*_1_ (0) = 1, *M*_2_ (0) = 0 and *E*_0_ = 0. For simulations, we sampled parameter values from a nonlinear mixed-effect model (*3*), with the following fixed effects and standard deviation of the random effects (in parenthesis): Log10 *β*: −7.23 (0.2) virions^-1^ day^-1^; *δ*: 3.13 (0.02) day^-1^ cells^-k^; *k*: 0.08 (0.02); Log10(*π*): 2.59 (0.05) day^-1^; *m*: 3.21 (0.33) days^-1^cells^-1^; Log10(*ω*): −4.55 (0.01) days^-1^cells^-1^. We also assumed *r* = 10; *δ*_*E*:_ = 1 day^-1^; *q* = 2.4 × 10^−5^ day^-1^ and *c* = 1*5* day^-1^.

### Intra-host transmission model

To estimate SARS-CoV-2 infectiousness *P*_*t*_ [*V*(*t*)] we employed the function, 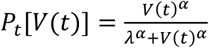. Here, *V*(*t*) is the viral load of the transmitter obtained from our previously proposed within host model and estimates (*2*). *λ* is the infectivity parameter that represents the viral load that corresponds to 50% infectiousness and 50% contagiousness, and *α* is the Hill coefficient that controls the slope of the dose-response curve. Our transmission model assumes that only some contacts of an infected individual with viral load dependent infectiousness are physically exposed to the virus (defined as exposure contacts), that only some exposure contacts have virus passaged to their airways (contagiousness) and that only some exposed contacts with virus in their airways become secondarily infected (successful secondary infection). Contagiousness and infectiousness are then treated as viral load dependent multiplicative probabilities with transmission risk for a single exposure contact being the product. Contagiousness is considered to be viral load dependent based on the concept that a transmitter’s dispersal cloud of virus is more likely to prove contagious at higher viral load, which is entirely separate from viral infectivity within the airway once a virus contacts the surface of susceptible cells.

We assumed that the total exposed contacts within a time step 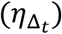 is gamma distributed, i.e. 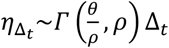, using the average daily contact rates (*θ*) and the dispersion parameter (*ρ*). To obtain the true number of exposure contacts with airway exposure to virus, we multiply the contagiousness of the transmitter by the total exposed contacts within a time step (i.e., 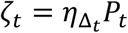). Transmissions within a time step are simulated stochastically using time-dependent viral load to determine infectiousness (*p*_*t*_). Successful transmission is modelled stochastically by drawing a random uniform variable (*U*(0,1)) and comparing it with infectiousness of the transmitter. In the case of successful transmission, the number of secondary infections within that time step 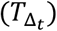 is obtained by the product of the infectiousness (*p*_*t*_) and the number of exposure contacts drawn from the gamma distribution (*ζ*_*t*_). In other words, the number of secondary infections for a time step is 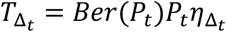. We obtain the number of secondary infections from a transmitter on a daily basis noting that viral load, and subsequent risk, does not change substantially within a day. We then summed up the number of secondary infections over 30 days since the time of exposure to obtain the individual reproduction number, i.e., 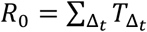.

We further assume that upon successful infection, it takes j days for the virus to move within-host, reach the infection site and produce the first infected cell. To calculate serial interval (time between the onset of symptoms of transmitter and secondarily infected person), we sample the incubation period in the transmitter and in the secondarily infected person from a gamma distribution (*4, 5*). In cases in which symptom onset in the newly infected person precedes symptom onset in the transmitter, the serial interval is negative; otherwise, serial interval is non-negative.

The model was fit to distributions of individual R0 (secondary transmissions per person) and serial interval as previously described (*6-10*). We then arrived at parameter estimates for *λ, τ, α* and *θ* and identified that a skewed distribution of daily exposure contacts explains the virus super-spreader property. This model was used to obtain baseline levels of secondary transmission for simulated placebo recipients.

**Supplementary figure 1.**
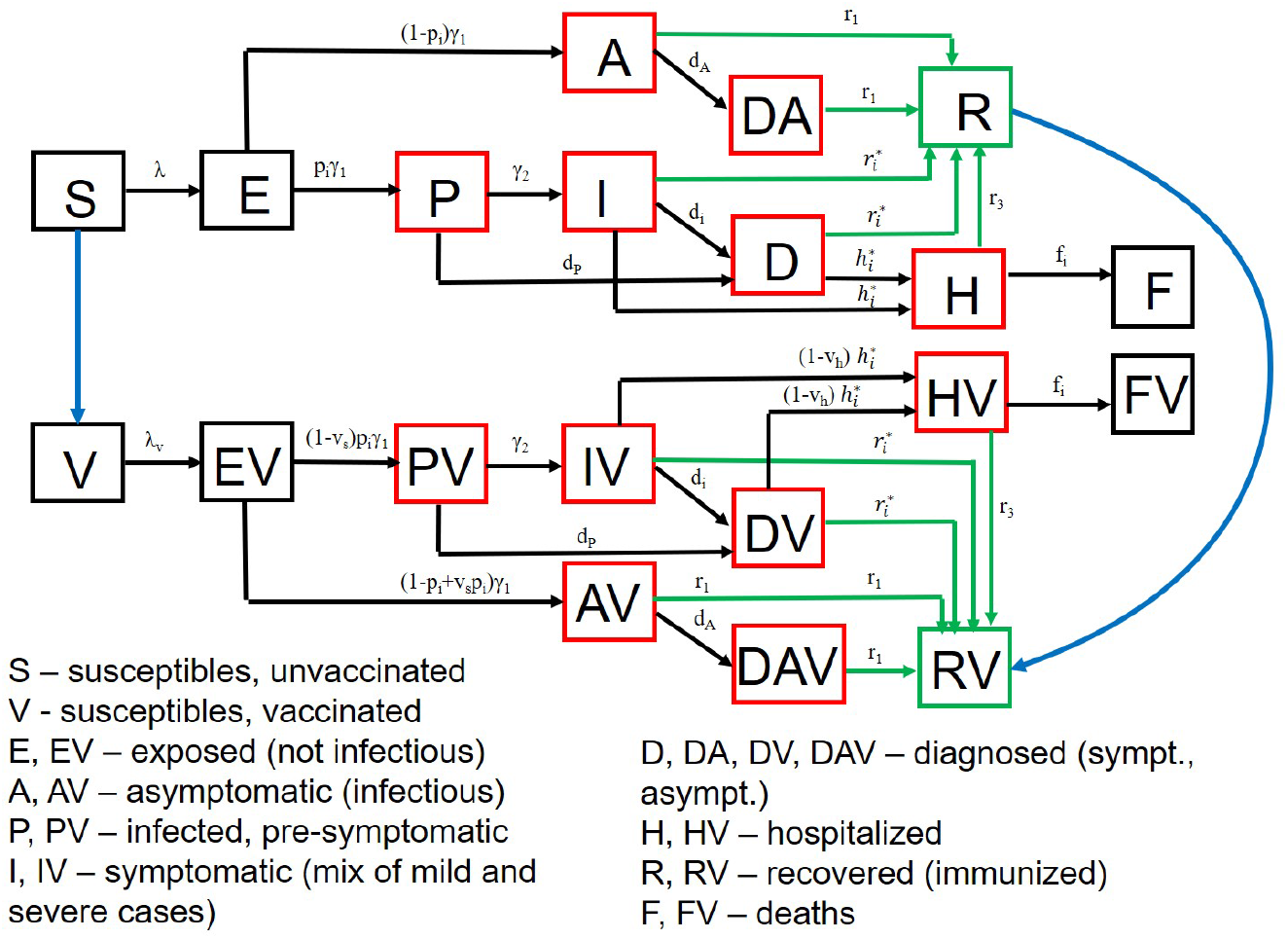
SARS-CoV-2 transmission model in King County, Washington. Model structure captures transition from susceptible (S) to exposed (E) to asymptomatic infection (A), or to pre-symptomatic (P) and then symptomatic infection (I) followed by recovery (R), hospitalization (H) or death (F). A similar potential pathway is also shown for a vaccinated cohort (V). Diagnosed (D) and diagnosed asymptomatic (DA) is an intermediate step for a proportion of people.

**Supplementary figure 2.**
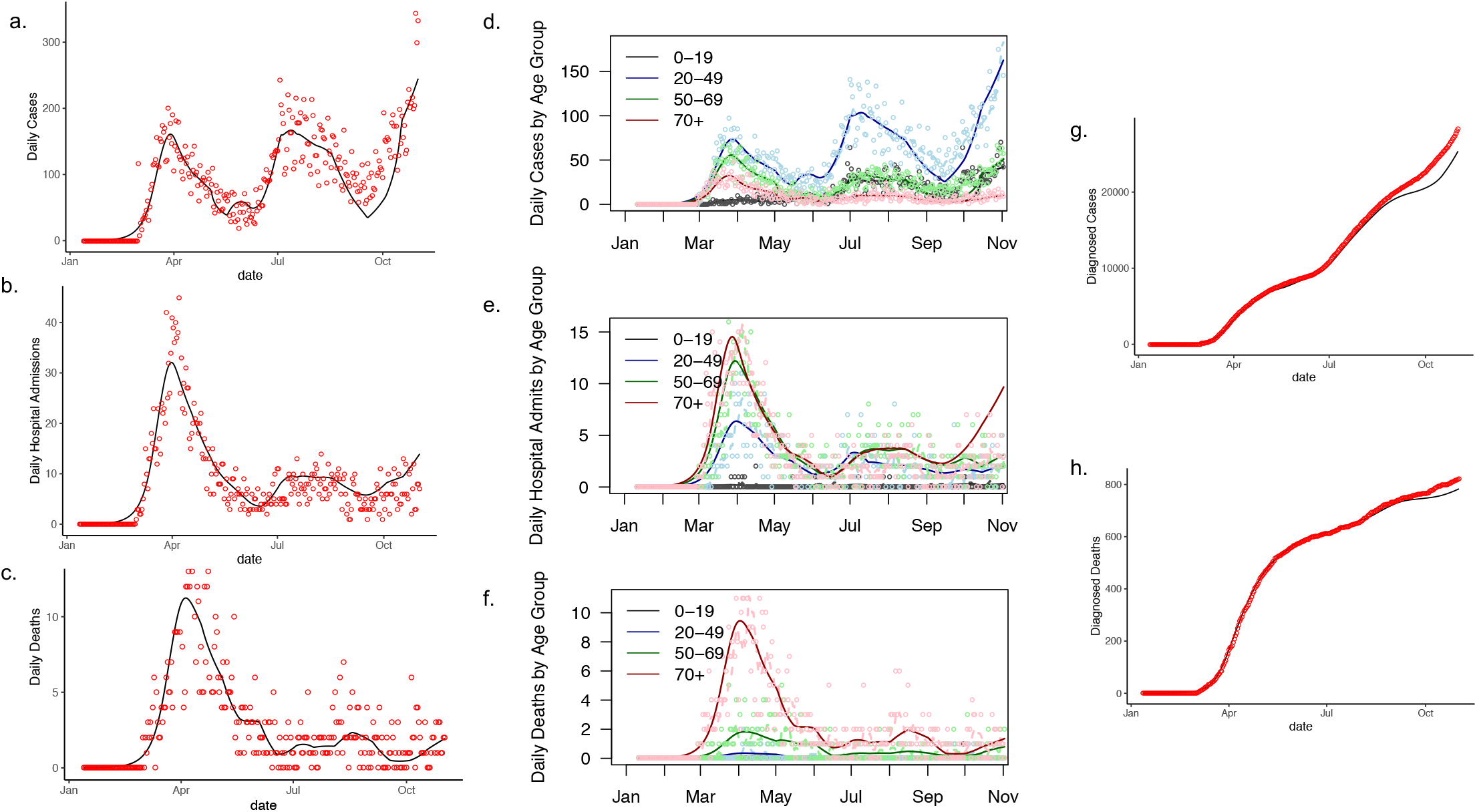
Calibration of a SARS-CoV-2 transmission model in King County, Washington between February and November 1, 2020. Model fit is to **a**. daily cases, **b**. daily hospitalizations, **c**. daily deaths, **d**. age-stratified cases, **e**. age-stratified hospitalizations, **f**. age-stratified deaths, **g**. cumulative cases, and **h**. cumulative deaths through the end of October 2021.

**Supplementary figure 3.**
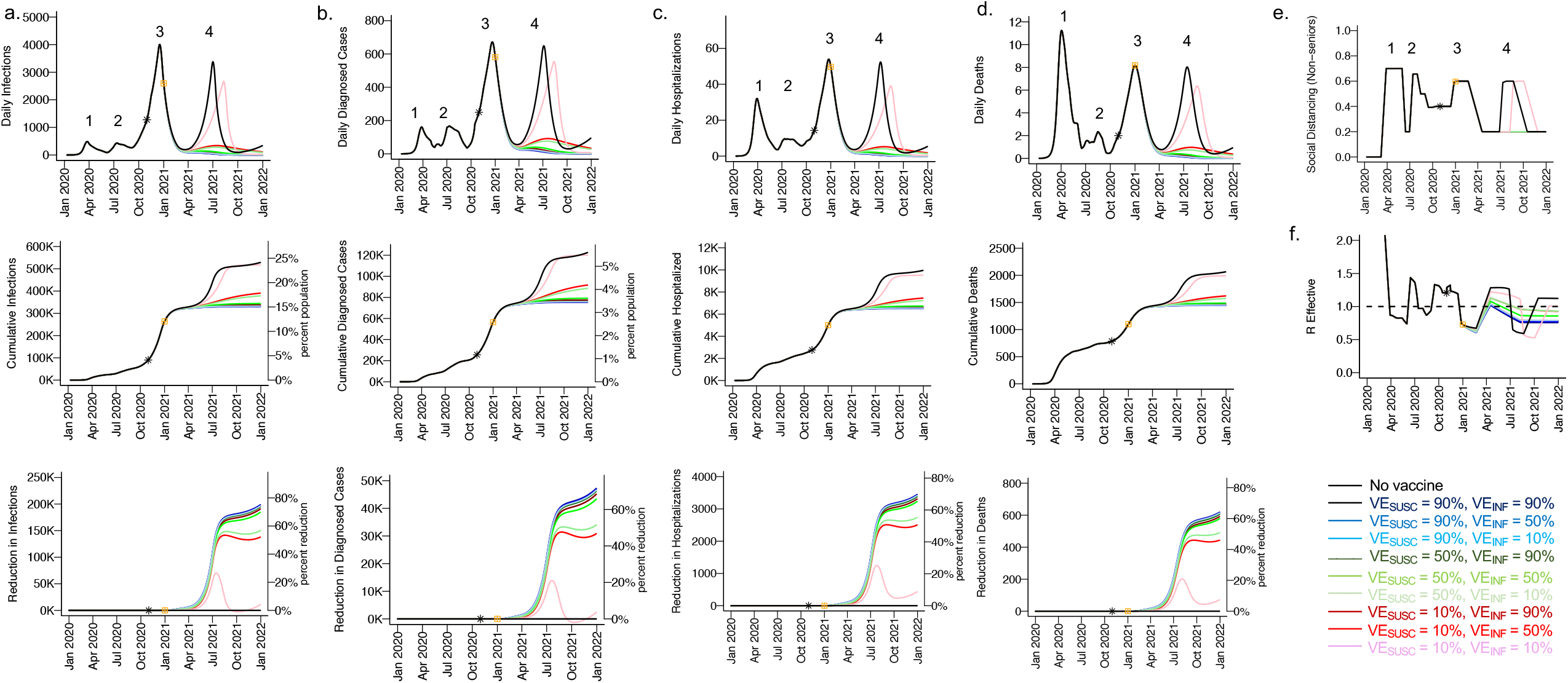
High VE_SUSC_ or high VE_INF_ alone can effectively limit cases and deaths with initial vaccine prioritization to the young and middle-aged. For unvaccinated (black) and each vaccine cohort (colored lines, legend), we project **a**. infections, **b**. diagnosed cases, **c**. hospitalizations and **d**. deaths, as well as **e**. social distancing relative to pre-pandemic s and **f**. the effective reproductive number. The first four columns **(a-d)** are organized by row: top = daily incidence, middle = cumulative, bottom = reduction since day of vaccination. Waves of infection are numbered 1-4. Nine combinations of VE_SUSC_ and VE_INF_ are considered while VE_SYMP_ is fixed at 10%. High VE_SUSC_ (90%) simulations are blue and have similar outcomes to one another. Moderate VE_SUSC_ (50%) simulations are green. Low VE_SUSC_ (10%) simulations are red / pink. Dark lines are high VE_INF_ (90%) and have similar outcomes to one her. Moderate darkness lines are medium VE_INF_ (50%). Light lines are low VE_INF_ (10%). The largest reduction in cases is associated with either high VE_SUSC_ or VE_INF_. 5000 vaccines are n per day starting January 1, 2021 (yellow square) until 50% are vaccinated.. Case threshold for reinstituting physical distancing to 0.6 is 300 per 100,000 and for relaxation is 100 per 80% of vaccines are initially allocated to the elderly with the remaining 20% to middle-aged cohorts.

**Supplementary figure 4.**
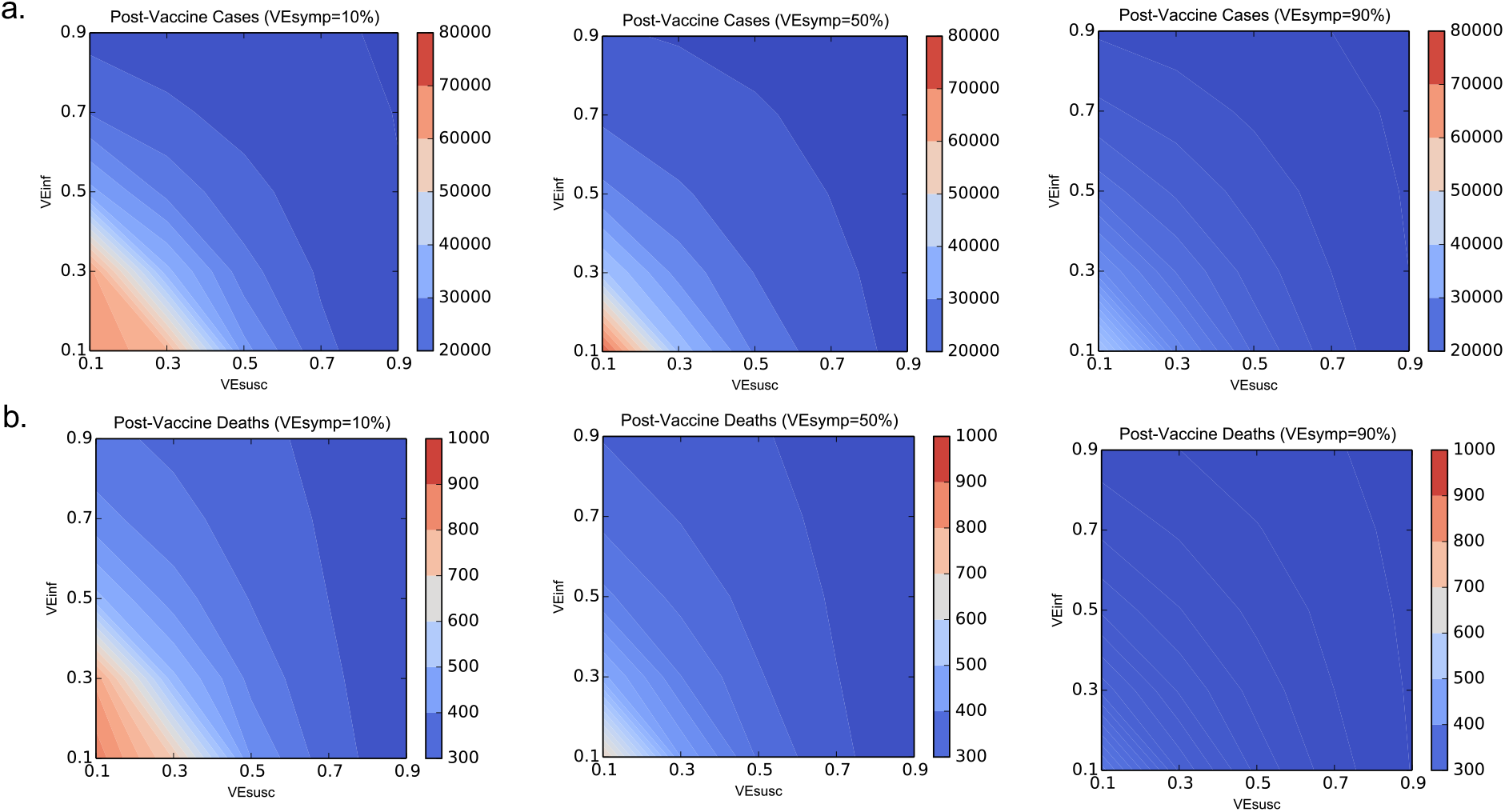
High VE_SUSC_ or high VE_INF_ independently drive reductions in SARS-CoV-2 cases and deaths. Heat maps comparing contrasting vaccine scenarios. **a**. Post-vaccine diagnosed cases (top row) and **b**. post-vaccine deaths (bottom row) with different combinations of VE_SUSC_ and VE_INF_. In this simulation, there were 62979 diagnosed cases and 1144 deaths prior to vaccination and heat maps capture all outcomes beyond this point. The left column assumes VE_SYMP_=10%; middle column assumes VE_SYMP_=50%; right column assumes VE_SYMP_=90%. VE_INF_ and VE_SUSC_ have similar impacts on both outcomes with benefit across a wide range of VE_SYMP_.

**Supplementary figure 5.**
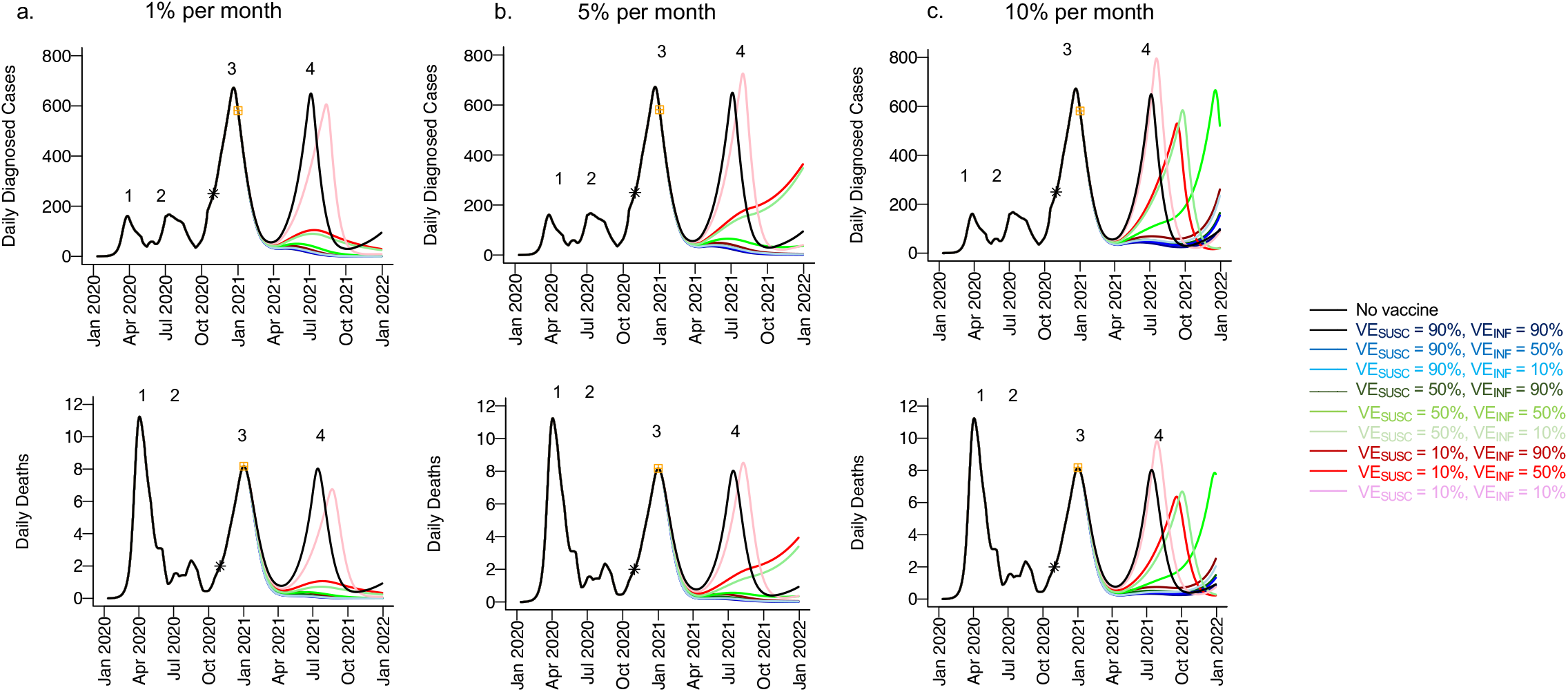
High VE_SUSC_ or high VE_INF_ most effectively limit cases and deaths in the context of waning vaccine efficacy. For unvaccinated (black lines) and each vaccine cohort (colored lines, legend), we project daily diagnosed cases (top row) and daily deaths (bottom row). Columns are **a**. 1% (left), **b**. 5% (middle) and **c**. 10% (right) of of persons with vaccine induced immunity reverting to susceptible per month. Waves of infection are numbered 1-4. Nine combinations of VE_SUSC_ and VE_INF_ are considered while VE_SYMP_ is fixed at 90%. High VE_SUSC_ (90%) simulations are blue and have similar outcomes to one another. Moderate VE_SUSC_ (50%) simulations are green. Low VE_SUSC_ (10%) simulations are red / pink. Dark lines are high VE_INF_ (90%) and have similar outcomes to one another. Moderate darkness lines are medium VE_INF_ (50%). Light lines are low VE_INF_ (10%). The largest reduction in cases is associated with either high VE_SUSC_ or VE_INF_ though slow rebound occurred with VE_SUSC_ or VE_INF_ 50%. 5000 vaccines are given per day starting January 1, 2021 (yellow square) until 50% are vaccinated. Case threshold for reinstituting physical distancing to 0.6 is 300 per 100,000 and for relaxation is 100 per 100,000. 80% of vaccines are initially allocated to the elderly with the remaining 20% to middle-aged cohorts.

**Supplementary figure 6.**
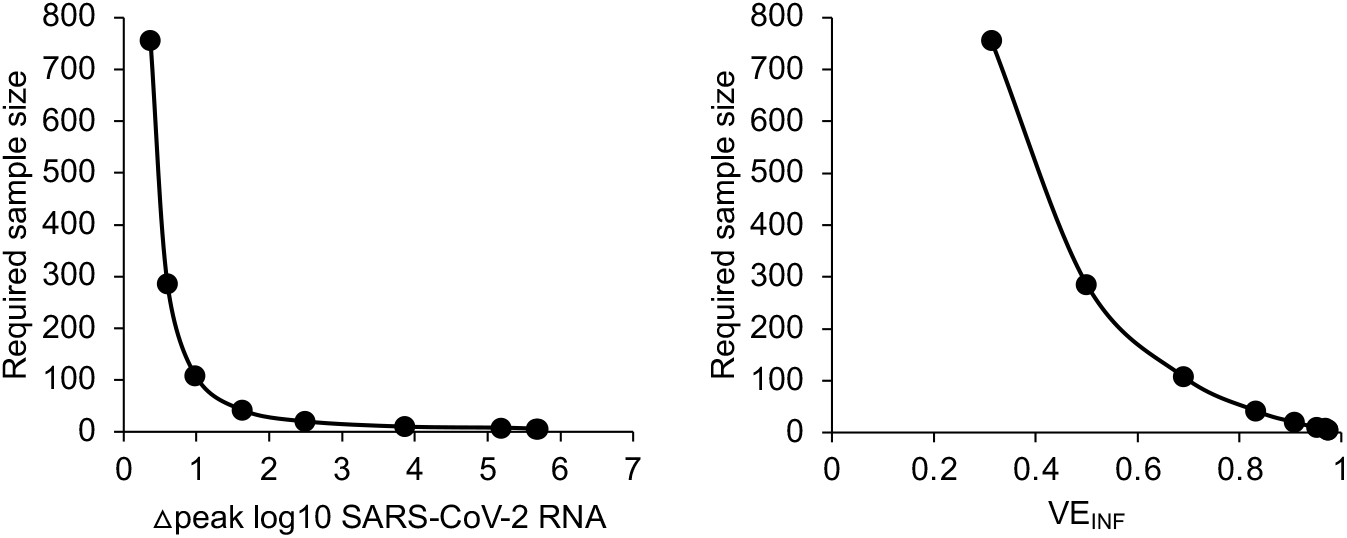
Required study size for human challenge studies to achieve 80% power. **a**. The relationship between mean peak viral load change and required sample size demonstrates that if a vaccine induces a >1.0 log_10_ reduction in peak viral load, then required sample size is much lower. **b**. The relationship between projected VE_I_ and required sample size demonstrates that if a vaccine induces VE_INF_>60%, then required sample size is much lower.

**Supplementary table 1.**
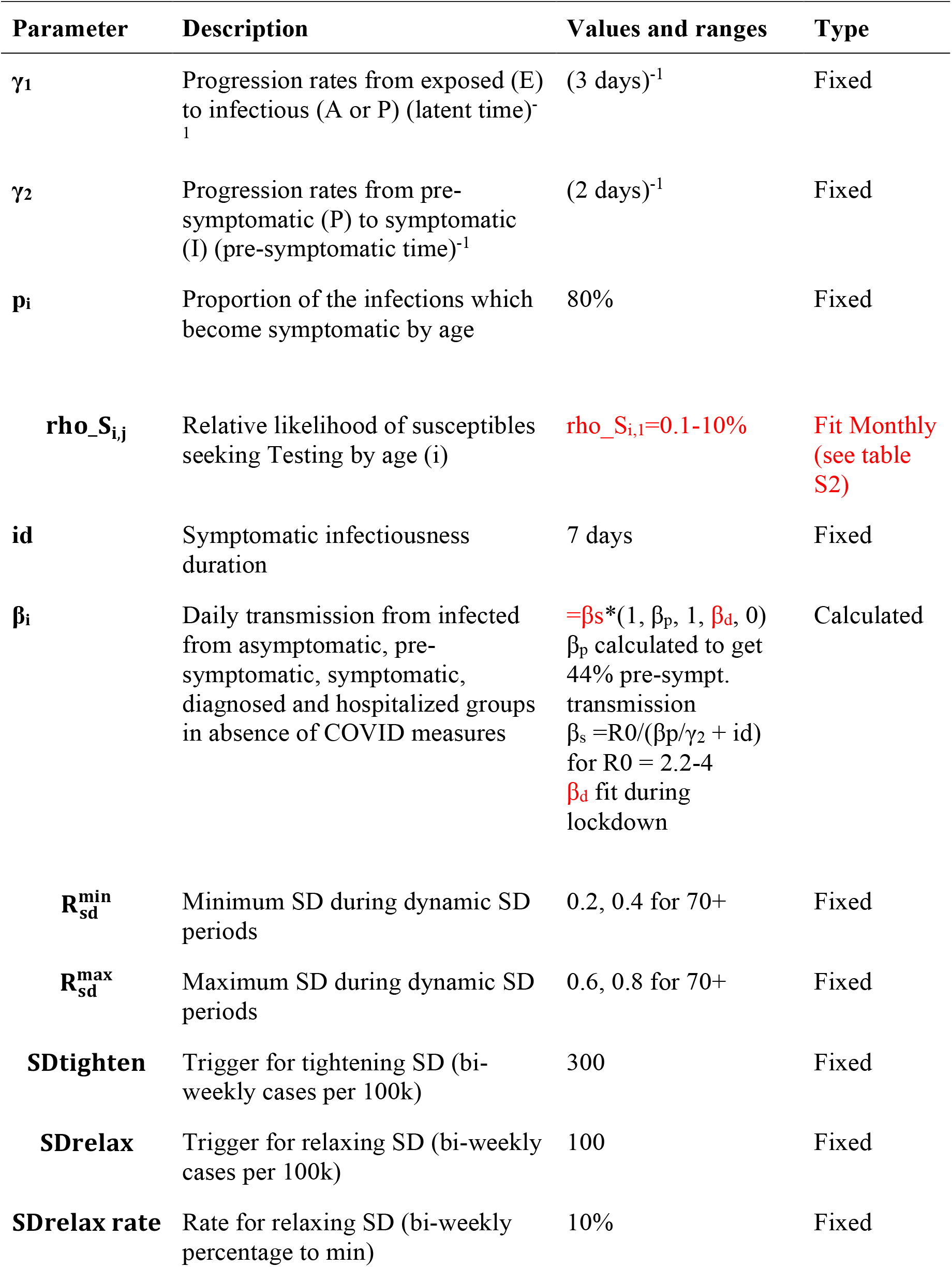
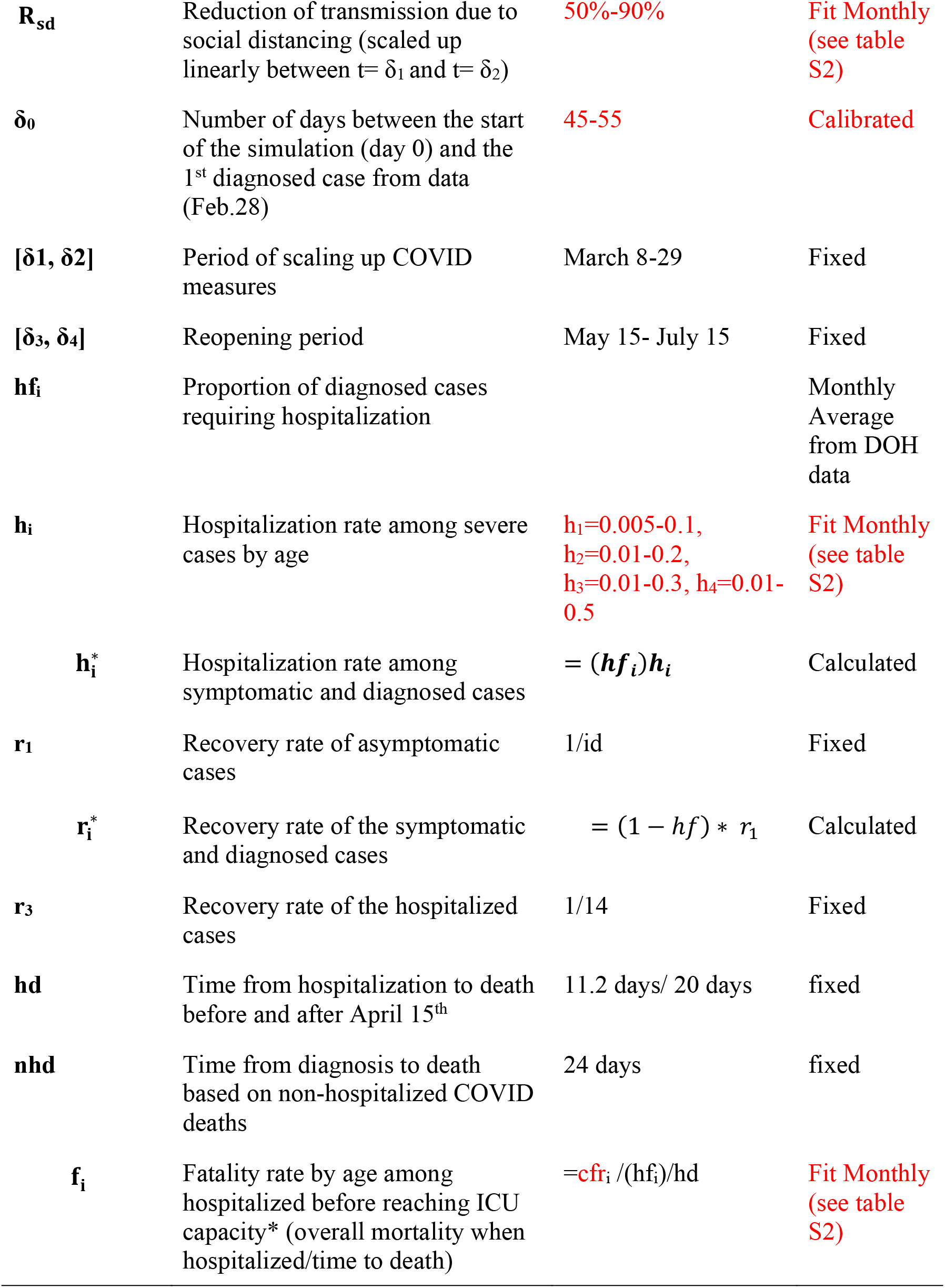
Parameters and ranges used in the analysis. (Fixed in black, Calibration in red)

| Parameter | Description | Values and ranges | Type |
| --- | --- | --- | --- |
| $\gamma_1$ | Progression rates from exposed (E) to infectious (A or P) (latent time)<sup>-1</sup> | (3 days)<sup>-1</sup> | Fixed |
| $\gamma_2$ | Progression rates from pre-symptomatic (P) to symptomatic (I) (pre-symptomatic time)<sup>-1</sup> | (2 days)<sup>-1</sup> | Fixed |
| $p_i$ | Proportion of the infections which become symptomatic by age | 80% | Fixed |
| $\rho_{S_{i,j}}$ | Relative likelihood of susceptibles seeking Testing by age (i) | $\rho_{S_{i,1}}=0.1-10\%$ | Fit Monthly (see table S2) |
| $id$ | Symptomatic infectiousness duration | 7 days | Fixed |
| $\beta_i$ | Daily transmission from infected from asymptomatic, pre-symptomatic, symptomatic, diagnosed and hospitalized groups in absence of COVID measures | $=\beta_s*(1, \beta_p, 1, \beta_d, 0)$<br>$\beta_p$ calculated to get 44% pre-sympt. transmission<br>$\beta_s = R_0/(\beta_p/\gamma_2 + id)$<br>for $R_0 = 2.2-4$<br>$\beta_d$ fit during lockdown | Calculated |
| $R_{sd}^{min}$ | Minimum SD during dynamic SD periods | 0.2, 0.4 for 70+ | Fixed |
| $R_{sd}^{max}$ | Maximum SD during dynamic SD periods | 0.6, 0.8 for 70+ | Fixed |
| **SDtighten** | Trigger for tightening SD (bi-weekly cases per 100k) | 300 | Fixed |
| **SDrelax** | Trigger for relaxing SD (bi-weekly cases per 100k) | 100 | Fixed |
| **SDrelax rate** | Rate for relaxing SD (bi-weekly percentage to min) | 10% | Fixed |
| $R_{sd}$ | Reduction of transmission due to social distancing (scaled up linearly between $t = \delta_1$ and $t = \delta_2$) | 50%-90% | Fit Monthly (see table S2) |
| $\delta_0$ | Number of days between the start of the simulation (day 0) and the 1<sup>st</sup> diagnosed case from data (Feb.28) | 45-55 | Calibrated |
| $[\delta_1, \delta_2]$ | Period of scaling up COVID measures | March 8-29 | Fixed |
| $[\delta_3, \delta_4]$ | Reopening period | May 15- July 15 | Fixed |
| $hf_i$ | Proportion of diagnosed cases requiring hospitalization | | Monthly Average from DOH data |
| $h_i$ | Hospitalization rate among severe cases by age | $h_1=0.005-0.1$,<br>$h_2=0.01-0.2$,<br>$h_3=0.01-0.3$, $h_4=0.01-0.5$ | Fit Monthly (see table S2) |
| $h_i^*$ | Hospitalization rate among symptomatic and diagnosed cases | $= (hf_i)h_i$ | Calculated |
| $r_1$ | Recovery rate of asymptomatic cases | 1/id | Fixed |
| $r_i^*$ | Recovery rate of the symptomatic and diagnosed cases | $= (1 - hf) * r_1$ | Calculated |
| $r_3$ | Recovery rate of the hospitalized cases | 1/14 | Fixed |
| $hd$ | Time from hospitalization to death before and after April 15<sup>th</sup> | 11.2 days/ 20 days | fixed |
| $nhd$ | Time from diagnosis to death based on non-hospitalized COVID deaths | 24 days | fixed |
| $f_i$ | Fatality rate by age among hospitalized before reaching ICU capacity* (overall mortality when hospitalized/time to death) | $= cfr_i / (hf_i) / hd$ | Fit Monthly (see table S2) |

**Supplementary table 2.** Monthly Parameter Fits. The columns represent the distribution of each parameter across the 4 age groups for each calibrated month:

| **Parameter** | **Age(y)** | **Lockdown** | **May** | **June** | **July** | **Aug** | **Sept** | **Oct** |
| --- | --- | --- | --- | --- | --- | --- | --- | --- |
| **rho_S<sub>i</sub>** | **0-19** | 0.026 | 0.016 | 0.016 | 0.041 | 0.033 | 0.034 | 0.003 |
|  | **20-49** | 0.008 | 0.014 | 0.017 | 0.034 | 0.035 | 0.03 | 0.023 |
|  | **50-69** | 0.006 | 0.025 | 0.04 | 0.053 | 0.046 | 0.036 | 0.012 |
|  | **70+** | 0.002 | 0.014 | 0.068 | 0.1 | 0.1 | 0.1 | 0.09 |
| **sd<sub>i</sub>** | **0-19** | 0.47 | 0.48 | 0 | 0.3 | 0.45 | 0.2 | 0.22 |
|  | **20-49** | 0.7 | 0.7 | 0.2 | 0.65 | 0.5 | 0.39 | 0.4 |
|  | **50-69** | 0.72 | 0.7 | 0.2 | 0.36 | 0.7 | 0.7 | 0.7 |
|  | **70+** | 0.9 | 0.9 | 0.4 | 0.9 | 0.9 | 0.9 | 0.9 |
| **h<sub>i</sub>** | **0-19** | 0.005 | 0.017 | 0.051 | 0.03 | 0.005 | 0.039 | 0.02 |
|  | **20-49** | 0.05 | 0.07 | 0.16 | 0.073 | 0.082 | 0.067 | 0.046 |
|  | **50-69** | 0.08 | 0.06 | 0.057 | 0.052 | 0.06 | 0.056 | 0.028 |
|  | **70+** | 0.14 | 0.12 | 0.057 | 0.05 | 0.05 | 0.05 | 0.05 |
| **cfr<sub>i</sub>** | **0-19** | 0 | 0 | 0 | 0 | 0 | 0 | 0 |
|  | **20-49** | 0.0004 | 0 | 0 | 0 | 0 | 0 | 0 |
|  | **50-69** | 0.034 | 0.007 | 0.005 | 0.005 | 0.006 | 0.005 | 0.005 |
|  | **70+** | 0.34 | 0.18 | 0.1 | 0.11 | 0.19 | 0.067 | 0.11 |

**Supplementary table 3.** Contact matrix. The columns represent the distribution of contacts of a person from given age group across all age groups:

| Proportion contacts with | 0-19 y | 20-49 y | 50-69 y | 70+ y |
| --- | --- | --- | --- | --- |
| 0-19 y | 0.56 | 0.24 | 0.15 | 0.18 |
| 20-49 y | 0.34 | 0.57 | 0.49 | 0.34 |
| 50-69 y | 0.08 | 0.16 | 0.29 | 0.28 |
| 70+ y | 0.01 | 0.03 | 0.07 | 0.20 |

**Supplementary table 4.** King County age pyramid based on data from 2017

| Proportion of the population | 0-19 y | 20-49 y | 50-69 y | 70+ y |
| --- | --- | --- | --- | --- |
|  | 22.93% | 45.52% | 23.50% | 8.05% |

**Supplementary table 5.**
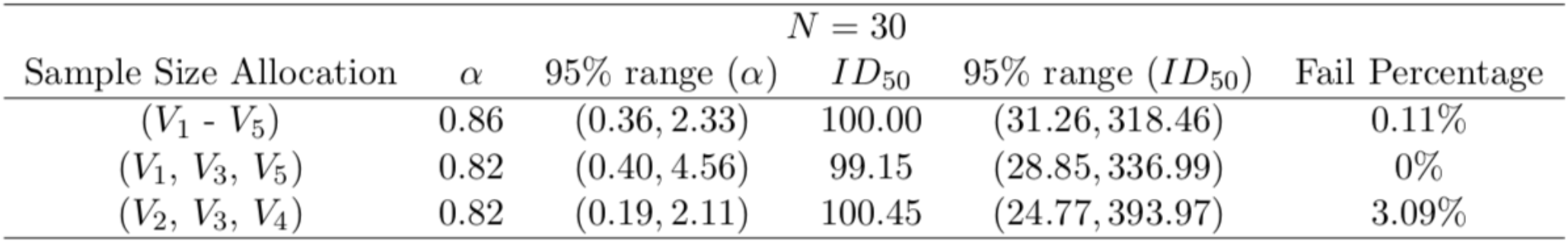
Simulation results for parameter estimation.

| $N = 30$ | | | | | | |
| --- | --- | --- | --- | --- | --- | --- |
| Sample Size Allocation | $\alpha$ | 95% range ($\alpha$) | $ID_{50}$ | 95% range ($ID_{50}$) | Fail Percentage | |
| $(V_1 - V_5)$ | 0.86 | (0.36, 2.33) | 100.00 | (31.26, 318.46) | 0.11% | |
| $(V_1, V_3, V_5)$ | 0.82 | (0.40, 4.56) | 99.15 | (28.85, 336.99) | 0% | |
| $(V_2, V_3, V_4)$ | 0.82 | (0.19, 2.11) | 100.45 | (24.77, 393.97) | 3.09% | |

**Supplementary table 6.** Simulation results for parameter estimation when ID50 is mis-specified.

| $N = 30$ | | | | | | |
| --- | --- | --- | --- | --- | --- | --- |
| $ID_{50}$ | $\alpha$ | 95% range ($\alpha$) | $ID_{50}$ | 95% range ($ID_{50}$) | Fail Percentage | |
| 100 pfu | 0.86 | (0.37, 2.33) | 100.00 | (31.26, 318.46) | 0.07% |  |
| 50 pfu | 0.87 | (0.37, 2.18) | 50.31 | (12.89, 153.19) | 0.12% |  |
| 10 pfu | 0.95 | (0.39, 6.45) | 10.78 | (2.92, 35.00) | 0.10% |  |
| 5 pfu | 0.99 | (0.38, 6.45) | 6.40 | (1.46, 20.41) | 0.14% |  |

**Supplementary table 7.** Simulation results for parameter estimation with flexible sample size allocation procedure.

| $ID_{50}$ | $\alpha$ | 95% range ($\alpha$) | $ID_{50}$ | 95% range ($ID_{50}$) | Fail Percentage | |
| --- | --- | --- | --- | --- | --- | --- |
| 50 pfu | 0.87 | (0.36, 2.18) | 50.31 | (12.32, 149.94) | 0.21% |  |
| 10 pfu | 0.87 | (0.28, 6.45) | 10.86 | (0.80, 38.92) | 2.75% |  |
| 5 pfu | 0.98 | (-0.21, 6.45) | 6.40 | (0.43, 17, 712.04) | 7.15% |  |

## References

1. D. V. Mehrotra et al., Clinical Endpoints for Evaluating Efficacy in COVID-19 Vaccine Trials. Ann Intern Med, (2020).

2. M. Lipsitch, N. E. Dean, Understanding COVID-19 vaccine efficacy. Science 370, 763–765 (2020).

3. FDA, in https://www.fda.gov/media/139638/download. (2020).

4. https://www.businesswire.com/news/home/20201116005608/en/. (2020).

5. https://www.businesswire.com/news/home/20201118005595/en/. (2020).

6. D. Bertsimas et al., Optimizing Vaccine Allocation to Combat the COVID-19 Pandemic. medRxiv, 2020.2011.2017.20233213 (2020).

7. J. H. Buckner, G. Chowell, M. R. Springborn, Dynamic Prioritization of COVID-19 Vaccines When Social Distancing is Limited for Essential Workers. medRxiv, 2020.2009.2022.20199174 (2020).

8. L. Matrajt, J. Eaton, T. Leung, E. R. Brown, Vaccine optimization for COVID-19: who to vaccinate first? medRxiv, 2020.2008.2014.20175257 (2020).

9. M. E. Halloran, M. Haber, I. M. Longini, C. J. Struchiner, Direct and indirect effects in vaccine efficacy and effectiveness. Am J Epidemiol 133, 323–331 (1991).

10. J. K. Yin et al., Systematic Review and Meta-analysis of Indirect Protection Afforded by Vaccinating Children Against Seasonal Influenza: Implications for Policy. Clin Infect Dis 65, 719–728 (2017).

11. R. I. Bailey et al., Pathogen transmission from vaccinated hosts can cause dose-dependent reduction in virulence. PLoS Biol 18, e3000619 (2020).

12. A. D. Paltiel, J. L. Schwartz, A. Zheng, R. P. Walensky, Clinical Outcomes Of A COVID-19 Vaccine: Implementation Over Efficacy. Health Aff (Millwood), 101377hlthaff202002054 (2020).

13. M. Voysey et al., Safety and efficacy of the ChAdOx1 nCoV-19 vaccine (AZD1222) against SARS-CoV-2: an interim analysis of four randomised controlled trials in Brazil, South Africa, and the UK. The Lancet.

14. A. Goyal, D. B. Reeves, E. F. Cardozo-Ojeda, J. T. Schiffer, B. T. Mayer, Wrong person, place and time: viral load and contact network structure predict SARS-CoV-2 transmission and super-spreading events. medRxiv, 2020.2008.2007.20169920 (2020).

15. L. Ferretti et al., Quantifying SARS-CoV-2 transmission suggests epidemic control with digital contact tracing. Science 368, eabb6936 (2020).

16. X. He et al., Temporal dynamics in viral shedding and transmissibility of COVID-19. Nat Med 26, 672–675 (2020).

17. Z. J. Madewell, Y. Yang, I. M. Longini, M. E. Halloran, N. E. Dean, Household transmission of SARS-CoV-2: a systematic review and meta-analysis of secondary attack rate. medRxiv, 2020.2007.2029.20164590 (2020).

18. M. E. Halloran et al., Simulations for designing and interpreting intervention trials in infectious diseases. BMC Med 15, 223 (2017).

19. J. Y. Kim et al., Viral Load Kinetics of SARS-CoV-2 Infection in First Two Patients in Korea. J Korean Med Sci 35, e86 (2020).

20. R. Wölfel et al., Virological assessment of hospitalized patients with COVID-2019. Nature, (2020).

21. A. Goyal, E. F. Cardozo-Ojeda, J. T. Schiffer, Potency and timing of antiviral therapy as determinants of duration of SARS-CoV-2 shedding and intensity of inflammatory response. Sci Adv 6, (2020).

22. J. Fajnzylber et al., SARS-CoV-2 viral load is associated with increased disease severity and mortality. Nat Commun 11, 5493 (2020).

23. T. Kirby, COVID-19 human challenge studies in the UK. The Lancet Respiratory Medicine 8, e96 (2020).

24. C. Bracis et al., Widespread testing, case isolation and contact tracing may allow safe school reopening with continued moderate physical distancing: A modeling analysis of King County, WA data. Infectious Disease Modelling 6, 24–35 (2021).

25. A. Goyal, E. F. Cardozo-Ojeda, J. T. Schiffer, Potency and timing of antiviral therapy as determinants of duration of SARS CoV-2 shedding and intensity of inflammatory response. medRxiv, 2020.2004.2010.20061325 (2020).

26. A. Goyal, D. B. Reeves, E. F. Cardozo Ojeda, B. T. Mayer, J. T. Schiffer, Slight reduction in SARS-CoV-2 exposure viral load due to masking results in a significant reduction in transmission with widespread implementation. medRxiv, 2020.2009.2013.20193508 (2020).

27. S. M. Kissler, C. Tedijanto, E. Goldstein, Y. H. Grad, M. Lipsitch, Projecting the transmission dynamics of SARS-CoV-2 through the postpandemic period. Science, (2020).

28. https://covid19-projections.com/us. (2020).

29. https://www.doh.wa.gov/Emergencies/COVID19/DataDashboard.

30. B. T. Mayer et al., Estimating the Risk of Human Herpesvirus 6 and Cytomegalovirus Transmission to Ugandan Infants from Viral Shedding in Saliva by Household Contacts. Viruses 12, (2020).

31. J. T. Schiffer, B. T. Mayer, Y. Fong, D. A. Swan, A. Wald, Herpes simplex virus-2 transmission probability estimates based on quantity of viral shedding. J R Soc Interface 11, 20140160 (2014).

32. A. Endo, S. Abbott, A. J. Kucharski, S. Funk, C. f. t. M. M. o. I. D. C.-W. Group, Estimating the overdispersion in COVID-19 transmission using outbreak sizes outside China. Wellcome Open Res 5, 67 (2020).

33. T. Ganyani et al., Estimating the generation interval for coronavirus disease (COVID-19) based on symptom onset data, March 2020. Euro Surveill 25, (2020).

34. J. T. Schiffer et al., Rapid localized spread and immunologic containment define Herpes simplex virus-2 reactivation in the human genital tract. Elife 2, e00288 (2013).

35. T. Watanabe, T. A. Bartrand, M. H. Weir, T. Omura, C. N. Haas, Development of a dose-response model for SARS coronavirus. Risk Anal 30, 1129–1138 (2010).

36. J. F. Chan et al., Simulation of the Clinical and Pathological Manifestations of Coronavirus Disease 2019 (COVID-19) in a Golden Syrian Hamster Model: Implications for Disease Pathogenesis and Transmissibility. Clin Infect Dis 71, 2428–2446 (2020).

37. S. F. Sia et al., Pathogenesis and transmission of SARS-CoV-2 in golden hamsters. Nature 583, 834–838 (2020).

38. F. J. Ibarrondo et al., Rapid Decay of Anti-SARS-CoV-2 Antibodies in Persons with Mild Covid-19. N Engl J Med 383, 1085–1087 (2020).

39. K. E. C. Ainslie, M. J. Haber, R. E. Malosh, J. G. Petrie, A. S. Monto, Maximum likelihood estimation of influenza vaccine effectiveness against transmission from the household and from the community. Stat Med 37, 970–982 (2018).

40. J. F. Seward, J. X. Zhang, T. J. Maupin, L. Mascola, A. O. Jumaan, Contagiousness of Varicella in Vaccinated CasesA Household Contact Study. JAMA 292, 704–708 (2004).

41. J. J. A. van Kampen et al., Shedding of infectious virus in hospitalized patients with coronavirus disease-2019 (COVID-19): duration and key determinants. medRxiv, 2020.2006.2008.20125310 (2020).

42. https://www.accessdata.fda.gov/scripts/cdrh/cfdocs/cfcfr/CFRSearch.cfm.

43. C. Bracis et al., Widespread testing, case isolation and contact tracing may allow safe school reopening with continued moderate physical distancing: a modeling analysis of King County, WA data. medRxiv, 2020.2008.2014.20174649 (2020).

44. D. Buitrago-Garcia et al., Occurrence and transmission potential of asymptomatic and presymptomatic SARS-CoV-2 infections: A living systematic review and meta-analysis. PLOS Medicine 17, e1003346 (2020).

45. Centers for Disease Control and Prevention, COVID-19 Pandemic Planning Scenarios.

46. Public Health - Seattle & King County, COVID-19 data dashboard. 2020.

47. MIDAS, Online Portal for COVID-19 Modeling Research. 2020.

48. N. M. Ferguson et al., Impact of non-pharmaceutical interventions (NPIs) to reduce COVID-19 mortality and healthcare demand.

49. S. M. Kissler et al., SARS-CoV-2 viral dynamics in acute infections. medRxiv, 2020.2010.2021.20217042 (2020).

## References

1. C. Bracis et al., Widespread testing, case isolation and contact tracing may allow safe school reopening with continued moderate physical distancing: A modeling analysis of King County, WA data. Infectious Disease Modelling 6, 24–35 (2021).

2. A. Goyal, E. F. Cardozo-Ojeda, J. T. Schiffer, Potency and timing of antiviral therapy as determinants of duration of SARS-CoV-2 shedding and intensity of inflammatory response. Sci Adv 6, (2020).

3. A. Goyal, D. B. Reeves, E. F. Cardozo-Ojeda, J. T. Schiffer, B. T. Mayer, Wrong person, place and time: viral load and contact network structure predict SARS-CoV-2 transmission and super-spreading events. medRxiv, 2020.2008.2007.20169920 (2020).

4. T. Ganyani et al., Estimating the generation interval for coronavirus disease (COVID-19) based on symptom onset data, March 2020. Euro Surveill 25, (2020).

5. S. A. Lauer et al., The Incubation Period of Coronavirus Disease 2019 (COVID-19) From Publicly Reported Confirmed Cases: Estimation and Application. Ann Intern Med 172, 577–582 (2020).

6. Q. Bi et al., Epidemiology and transmission of COVID-19 in 391 cases and 1286 of their close contacts in Shenzhen, China: a retrospective cohort study. Lancet Infect Dis, (2020).

7. Y. Zhang, Y. Li, L. Wang, M. Li, X. Zhou, Evaluating Transmission Heterogeneity and Super-Spreading Event of COVID-19 in a Metropolis of China. Int J Environ Res Public Health 17, (2020).

8. A. Dillon et al., Clustering and superspreading potential of severe acute respiratory syndrome coronavirus 2 (SARS-CoV-2) infections in Hong Kong. PREPRINT (Version 1) available at Research Square, (2020).

9. A. Endo, Centre for the Mathematical Modelling of Infectious Diseases COVID-19 Working Group,S. Abbott, A. Kucharski, S. Funk, Estimating the overdispersion in COVID-19 transmission using outbreak sizes outside China. Wellcome Open Res 5, (2020).

10. Z. Du et al., Serial Interval of COVID-19 among Publicly Reported Confirmed Cases. Emerg Infect Dis 26, 1341–1343 (2020).

